# Phylogenomics and population genomics of SARS-CoV-2 in Mexico during the pre-vaccination stage reveals variants of interest B.1.1.28.4, B.1.1.222 or B.1.1.519 and B.1.243 with mutations in the Spike protein and the Nucleocapsid

**DOI:** 10.1101/2021.05.18.21256128

**Authors:** Francisco Barona-Gómez, Luis Delaye, Erik Díaz-Valenzuela, Fabien Plisson, Arely Cruz-Pérez, Mauricio Díaz-Sánchez, Christian A. García-Sepúlveda, Alejandro Sanchez-Flores, Rafael Pérez-Abreu, Francisco J. Valencia-Valdespino, Natali Vega-Magaña, José Francisco Muñoz-Valle, Octavio Patricio García-González, Sofía Bernal-Silva, Andreu Comas-García, Angélica Cibrián-Jaramillo

## Abstract

Understanding the evolution of SARS-CoV-2 virus in various regions of the world during the Covid19 pandemic is essential to help mitigate the effects of this devastating disease. We describe the phylogenomic and population genetic patterns of the virus in Mexico during the pre-vaccination stage, including asymptomatic carriers. A RT-qPCR screening and phylogenomics reconstructions directed a sequence/structure analysis of the Spike glycoprotein, revealing mutation of concern E484K in genomes from central Mexico, in addition to the nationwide prevalence of the imported variant 20C/S:452R (B.1.427/9). Overall, the detected variants in Mexico show Spike protein mutations in the N-terminal domain (i.e., R190M), in the receptor-binding motif (i.e., T478K, E484K), within the S1-S2 subdomains (i.e., P681R/H, T732A), and at the basis of the protein, V1176F, raising concerns about the lack of phenotypic and clinical data available for the variants of interest (VOI) we postulate: 20B/478K.V1 (B.1.1.222 or B.1.1.519) and 20B/P.4 (B.1.1.28.4). Moreover, the population patterns of Single Nucleotide Variants (SNVs) from symptomatic and asymptomatic carriers obtained with a self-sampling scheme confirmed the presence of several fixed variants, and differences in allelic frequencies among localities. We identified the mutation N:S194L of the Nucleocapsid protein associated with symptomatic patients. Phylogenetically, this mutation is frequent in Mexican sub-clades, so we propose an additional VOI, 20A/N:194L.V2 (B.1.243). Our results highlight the dual and complementary role of Spike and Nucleocapsid proteins in adaptive evolution of SARS-CoV-2 to their hosts and provide a baseline for specific follow-up of mutations of concern during the vaccination stage.

**IMPACT STATEMENT:** Following self-sampling, screening of mutations of concern, and a combined phylogenomic and population genetics pipeline, we reveal the appearance of three VOI with mutations in the Spike protein, P.4 (B.1.1.28.4) and 20B/478K.V1 (B.1.1.222, leading to B.1.1.519), and in the Nucleocapsid protein, 20A/N:194L.V2 (B.1.243), in Mexico during the pre-vaccination stage. The mutation S194L in the Nucleocapsid was found to associate with symptomatic patients versus asymptomatic carriers in the population investigated. Our research can aid epidemiological genomics efforts during the vaccination stage in Mexico by contributing with a combined analytical platform and information about variants within different genetic lineages with the potential to evolve into variants of concern (VOC).

## INTRODUCTION

Mutation D614G of SARS-CoV-2 Spike protein was the first mutation implicated in increased transmission and a more efficient viral replication in human cells (Korber et al. 2020). It emerged as a consequence of natural selection in the S1 region of the Spike protein as the virus colonised human populations outside of Asia to Europe and to the rest of the world (Zhang et al. 2020). Shortly after the appearance of D614G, other mutations in the Receptor Binding Domain (RBD) of the Spike protein appeared, including but not limited to K417N, L452R, E484K and N501Y. Their increasingly high frequency in recent months, their potential for increased transmission coupled with immunological escape (Greaney et al. 2021; Li et al. 2020, 2021; Zhou et al. 2021; Garcia-Beltran et al. 2021), and the likely increased mortality (Davies et al. 2021) have alerted the medical and scientific community to follow emerging and prevalent variants as Variants of Interest (VOI) and Variants of Concern (VOC) (“WHO | SARS-CoV-2 Variants” 2021; Scientific Advisory Group for Emergencies 2021). VOI and VOC are both defined as highly occurring mutations and/or providing an increased fitness to the virus with a concomitant phenotypic change that results in an increased threat to human health (Resende et al. 2021), but VOI require experimental evidence of the suspected phenotypic dangerous trait to become VOC.

Emerging variants can be detected by their phylogenetic position provided by the Nextstrain (Hadfield et al. 2018) and Pangolin platforms, the latter being the primary source of classification and nomenclature of SARS-CoV-2 (Rambaut et al. 2020). Mutations included in VOI and VOC can also co-occur with other mutations, both within the variable N-terminal domain (NTD) region and/or the flexible receptor-binding domain (RBD) of the Spike protein or throughout the SARS-CoV-2 genome. Such co-occurring mutations, including indels, are expected given the virus’ current evolutionary trajectory. Of the sixteen emerging lineages identified during the summer of 2020, four evolved early in 2021 into lineages with VOCs bearing signature shared mutations: 20I/501Y.V1 (United Kingdom, B.1.1.7, alpha), 20H/501Y.V2 (The Republic of South Africa, B.1.351, beta), and 20J/501Y.V3 (Brazil, P.1 or B.1.1.28.1, gamma); and 20C/S:452R (California USA, B.1.427/9, epsilon). Together with the P.1 closely related VOI P.2 (B.1.1.28.2, zeta) these VOC/VOI often share mutations E484K and/or N501Y. Mutations defining VOC, e.g. E484K, are increasingly common worldwide as they are imported from their regions of origin into naive populations; but also as independent evolution converges to the same point mutations, for instance, in the recently reported VOI 20A/S.484K and 20C/S.484K (B.1.525/526, eta/iota, Lasek-Nesselquist et al. 2021; Annavajhala et al. 2021), amongst others.

These viral evolutionary emerging scenarios are worrisome for their potential to extend the duration of the pandemic, as these lineages emerge and spread in non-vaccinated populations concomitantly to vaccinated ones. The convergence of K417N, E484K and N501Y in different geographical regions suggest that during the “pre-vaccination stage” of the pandemic, similar selection pressures took place to increase SARS-CoV-2’s fitness (McCormick et al. 2021). For instance, 20J/501Y.V3 or P.1 evolved in parallel with the 20B/S.484K or P.2 variant in Brazil, and they share the Spike protein mutations E484K and V1176F, but not N501Y. Likewise, although geographically unrelated, it can be argued that the places of origin of VOC 20I/501Y.V1, 20H/501Y.V2, 20J/501Y.V3 and 20C/S:452R have in large common populations with worrying recorded epidemiological statistics (e.g. deaths, cases, R0, positivity, etc.). Cases where natural or unintended immunological pressures, such as the use of convalescent plasma (Kemp et al. 2021) or high prevalence of the disease leading to a high immunological burden (Resende et al. 2021), can help us understand what to expect during the vaccination stage of the pandemic, but further population genomics and epidemiological studies are needed.

The finer-scale population genetic patterns of the virus as it adapts to local conditions is a recent focus of research. SARS-CoV-2 has a high-fidelity transcription and replication process (Ma et al. 2015) compared to other single-stranded RNA viruses, resulting in generally lower genetic diversity (Rausch et al. 2020). Yet the large SARS-CoV genomes allow efficient exploration of the sequence space (Forni et al. 2017) so the array of possible variants during SARS-CoV-2’s trajectory as it adapts to humans’ large populations and even crosses over to other species (Oude Munnink et al. 2021) is yet to be described in full. One of the few previous population-level studies in SARS-CoV-2 showed that haplotype diversity and allele frequencies in infection clusters are likely caused by natural selection (Liu et al. 2020). Additionally, in early 2020 there were two significant lineages identified by population-level analyses (Tang et al. 2020); in late 2020, there were six viral subtypes based on the estimation of increased nucleotide diversity (Morais et al. 2020; Yang et al. 2020); and in 2021, there were four sub-strains in the US alone detected by population-level analyses (Wang et al. 2021).

Here, we provide a baseline analysis for the evolution of SARS-CoV-2 in Mexico between March 2020, when the first case of Covid19 was registered in the country, and February 2021, when large-scale vaccination started. Based on such analysis, we postulate the occurrence of two VOI: P.4, which evolved from 20B/P.2, and 20B/478K.V1 or B.1.1.222 / B.1.1.519, emerging within Mexico. We use publicly available genomic data for Mexico, plus 85 genomes generated by us that include asymptomatic carriers identified using a novel self-sampling strategy developed to increase success rate in sampling these individuals without uncertainty and reduced risk. Two specific SARS-CoV-2 scenarios, the prevalence of mutations of concern and population-level viral allelic variation, are reported, while providing an *ad hoc* bioinformatics pipeline for real-time analysis of SARS-CoV-2 genomes in Mexico. Our report is timely as it coincides with a national epidemiological genomics program that aims at providing a thousand genomes per month from hospital samples throughout the country; and of interest as the current vaccination program in many countries including Mexico, involves diverse vaccine technologies.

## METHODS

### Self-sampling for identification of asymptomatic SARS-CoV-2 carriers and sample preparation

We sampled asymptomatic SARS-CoV-2 carriers from private organizations represented by a Human Resources (HR) in the City of Irapuato, State of Guanajuato. The HR contact person would report on suspected cases of Covid19 with symptoms, which were sent to the clinic for an assessment and diagnosis by health care professionals. At this point, all contacts of the suspected Covid19 patient were identified through contact tracing, including their family members, and provided with a self-sampling kit with instructions for collection of nasopharyngeal swabs (**Figure S1**). Validation in 58 volunteers was done by self-sampling and assisted sampling by a qualified healthcare professional, followed by quantification of nucleic acids with Qubit™ Flex Fluorometer (Thermo Fisher), and by qPCR amplification of the human marker RNAse P as control (**Table S1**).

Additive manufacturing technology became an essential player as supply shortages affected swab and viral transport media (VTM) production globally. Our group used 3D printing for emergency manufacturing devices, including face shields and swabs. For this, we partnered with Prothesia (https://www.prothesia.com/covid19), a Mexico based company dedicated to creating software algorithms to manufacture custom orthotic devices. FDA-cleared 3D-printed NP swabs designed by USF-Northwell and manufactured by Prothesia were used for self-sampling. Manufacturing was done using methacrylate monomers, urethane dimethacrylate and a photoinitiator as reagents, with stereolithography technology on Formlabs printers, curing and washing stations. The tip of the swab features a biomimetic design based on the “cattail” (Typha) plant, and it has a rounded nose to maximise comfort and lateral alternating nubs to maximize surface area for sample collection.

Samples were transported and saved in 700 µl of VTM (Atila Biosystems Inc) and/or DNA/RNA Shield (Zymo Research). The DNA was saved for future investigations. This provided enough material for two extractions of RNA, each of 300 µl, done with chemagic™ Viral DNA/RNA 300 Kit H96 from PerkinElmer, Quick-RNA Viral Kit (Zymo Research) or QIAmp Viral RNA Mini Kit (Qiagen); and one extraction of DNA of the entire 700 µl, done with ZymoBiomics DNA Kit. Around 31.5 ng/µl of RNA and 73.3 ng/µl of DNA were obtained per extraction in a total volume of circa 50 µl. The RNA material was used for SARS-CoV-2 diagnostics using suitable thermocyclers for RT-qPCR, with kits validated by InDRE and provided by Genes2Life, Mexico, to the academic diagnostics laboratories of Guanajuato (Wov19) and Jalisco (Decov triplex). Detection of San Luis Potosí SARS-CoV2 positive samples was done with the GeneFinder COVID-19 PLUS RealAmp Kit.

### Identification of mutations of concern

Screening for E484K and N501Y mutations was done by RT-qPCR, using module 2 of the MUT-SARS-CoV-2 kit (Genes2Life SAPI de CV, Mexico) simultaneously in the same reaction using RNA previously identified as positive for SARS-CoV-2, as previously (Vega-Magaña et al. 2021). The reactions were carried out using a QuantStudio 5 RT System thermal cycler from Applied Biosystem. This assay uses four fluorescent probes that specifically hybridize with the target sequences to discriminate each base/mutation. Thus, the detection and discrimination are carried out in a fourplex assay, being the channels for the FAM and HEX dyes dedicated to detection of wildtype background; and the Cal Fluor Red 610 and Quasar 670 dyes channels used for detection of the E484K and N501Y mutations, respectively. A total of 1560 RNA samples from the States of San Luis Potosí (170), Jalisco (1330) and Mexico City (60), previously identified as positive for SARS-CoV-2, were screened.

### Genome assembly and sequencing of SARS-CoV-2

Two different approaches were used to produce sequencing libraries. A total of 78 cDNA samples from Guanajuato (45) and San Luis Potosi (33) were prepared using the Illumina/IDbyDNA Respiratory Pathogen ID/AMR Enrichment Kit following the vendor’s protocol for library preparation. Libraries were prepared with Illumina RNA Prep with Enrichment (Illumina, Catalog no. 20040536) and IDT for Illumina DNA/RNA UD Indexes (Illumina, Catalog no. 20027213). Illumina RNA Prep with Enrichment consists in a On-Bead Tagmentation followed by a single hybridization step to generate enriched DNA and RNA libraries. After amplification, libraries were enriched as 3-plex reactions using the Illumina Respiratory Pathogen ID/AMR Panel to detect several viral (RNA) and bacterial (DNA) pathogens, including SARS-CoV-2, via probe capture. The DNA bacterial data will be reported elsewhere. Libraries were sequenced in a Illumina NextSeq 500 platform using a configuration for 75bp paired-end. Further 24 sequencing libraries from Jalisco (9) and San Luis Potosí (15) were constructed using the QIAGEN library preparation kit QIAseq_SARSCov2_Primer_Panel. Shotgun sequencing was done in a Illumina MiSeq 2×150 v3 platform. SARS-CoV-2 genome sequences were generated by mapping reads to the NC_045512.2 reference genome using the Explify pipeline at Illumina BaseSpace hub site or with Illumina DRAGEN Bio-IT Platform. Of the 102 genomic libraries generated only 35 reached GISAID quality for the phylogenomics analysis and 85 were suitable for the population genomics. Details on these genome sequences are provided in supplementary **Table S2**.

### Phylogenomics of SARS-CoV-2 mutations of concern

All complete and high-coverage SARS-CoV-2 genome sequences from Mexico (from samples obtained up to February 2021) were downloaded from GISAID in March 2021 (https://www.gisaid.org/). These were 1554 genomes. We also downloaded from GISAID all genome sequences represented in Nextstrain SARS-CoV-2 global analysis (https://nextstrain.org/ncov/global, date: March 2021). We joined the above set of sequences (and their associated metadata) into a single set for phylogenomic analyses by using in-house Perl scripts (https://github.com/luisdelaye/Mexstrain), giving place to our *ad hoc* Mexstrain platform (http://www.ira.cinvestav.mx/ncov.evol.mex.aspx) for phylogenomic analyses using a local version of Nextstrain (Hadfield et al. 2018). We used a ‘global’ sampling scheme grouping 10 sequences per ‘country year month’ and modified the defaults/include.txt file to include all Mexican sequences. For the rest of the parameters we used those provided by the ncov Nextstrain build. All configuration files are available upon request. The obtained phylogenomics were used to manually investigate mutations of concern according to Pangolin nomenclature (Rambaut et al. 2020).

### Incidences and relationships between SARS-CoV-2 mutations of concern

We focused on the gene coding for the Spike protein. All 1554 Mexican SARS-CoV-2 sequences were compared against MN908947 as reference used by Nextstrain (the sequence MN908947 used by Nextstrain version 3.0.3 is identical to NC_045512.2). We identified 337 combinations of mutations and 315 unique mutations from 1552 sequences using in-house Perl and Python scripts (two sequences were filtered out because of quality issues). To identify homologous positions between Mexican and reference sequences, we used the multiple sequence alignment generated by our local Nextstrain installation. We transformed the output file to study the incidence for 315 mutations grouped in 11 clades, and we studied their covariances between one another using in-house scripts with R packages: tidyverse (Wickham et al. 2019), circlize (Gu et al. 2014), and Python modules: NumPy (Harris et al. 2020), Pandas (McKinney et al. 2010), matplotlib (Hunter, 2007), seaborn (Waskom et al. 2017) - R version 4.0.4 (Ripley 2001), R Studio 1.4.1106 (“RStudio”), Python 3.8 (Van Rossum & Drake 2009), and JupyterNotebook 6.1.4 (Kluyver et al. 2016) (https://github.com/plissonf/Phylogenomics_SARS-CoV-2_Mexico).

### Population genomics of SARS-CoV-2

Nucleotide variants and small indels allow the assessment of allele frequencies at the population level and the intra-host co-occurrence of ancestral (known Wuhan reference) and derived (alternative) alleles that can inform the appearance of new variants and their evolution. We designed a mapping-based approach to call SNVs and indels directly from sequencing reads rather than assembled genomes. Due to the possibility of having SNPs due to RNA modification rather than RNA replication errors, we did not separate allele frequencies (i.e. A–G and C–T mismatches) that could potentially be due to deamination events (Li et al. 2020), so we refer to nucleotide variation at the population level as Single Nucleotide Variants (SNVs) more generally. Also, most CT values ranged between 28 and 30, suggesting an intermediate to high viral load in our samples (Zacharioudakis et al. 2021), reducing the possibility of biases in allele frequencies due to lower viral loads (Lythgoe et al. 2021). Raw reads from a total number of 102 pair-ended libraries were processed by first removing adapters, low quality bases and short reads using the fastP program (Chen et al. 2018) using default parameters. The reads were mapped to the NC_045512.2 version of the SARS-CoV-2 reference genome using BWA (Li & Durbin 2009) with default parameters. Sam alignments were converted to bam files and sorted using samtools (Li et al. 2009). Bam files were used to call variants (SNVs + indels) via the freebayes program (Garrison & Marth 2012). VCF files were then concatenated and parsed to retain only biallelic SNVs supported by at least 20 sequencing reads and alignment qualities above 30 using in-house R scripts. Allelic frequencies were estimated on the basis of 82 samples that passed our filters. Additionally, we used ANGSD (Korneliussen et al. 2014) to construct a genetic covariance matrix between samples using genotype likelihoods directly from the bam files, and used this matrix for a Principal Components Analysis using the prcomp function of R. To explore the distribution of SNVs within hosts, we identified and estimated the ‘allelic imbalance’, defined as the proportions of total reads that support either the ancestral or the derived allele, per polymorphic biallelic site. The statistical association of the allelic imbalance within hosts to the expression of symptoms was first tested using a Wilcoxon test collapsing all polymorphic sites within groups. A one-way ANOVA was then employed to test the individual effects of allelic imbalance at each polymorphic site to the expression of symptoms. Data visualization and statistical analysis were performed using the tidyverse (Wickham et al. 2019), ggridges (Wilke 2021) and rstatix (Kassambara 2021) R libraries.

### Positions of mutations of concern in the Spike protein of SARS-CoV-2

We extracted the Electron Microscopy structure (7A94, Benton et al. 2020) of the trimeric Spike (S) glycoprotein of SARS-CoV-2 bound to one Angiotensin-Converting Enzyme-2 (ACE2). We depicted different domains and subdomains of S in various colors and textures (cartoon, surfaces) using MacPyMOL v.1.7 (Schrodinger LLC, https://pymol.org/). We rendered 3D representations with ray before proceeding to screen captures. We marked the important mutations S477N, T478K, E484K, D614G, P681H/R and T732A.

### Ethical committee clearance

All the RNA samples that are reported in this study were obtained during a surveillance program part of a research protocol compliant with *El Comité de Bioética para la Investigación en Seres Humanos del Centro de Investigación y Estudios Avanzados del IPN* (COBISH) led by Dr Betzabe Quintanilla Vega, who approved research for all SARS-CoV-2 carriers in the State of Guanajuato (069/2020). The samples from the State of San Luis Potosi were obtained through the clinical diagnostic service provided by the *Centro de Investigación en Ciencias de la Salud y Biomedicina of the Universidad Autónoma de San Luis Potosi* as part of a research protocol compliant with the corresponding local authorities, represented by Dr Liliana Rangel Martinez according to the Ley Estatal de Salud and NOM-012-SSA3-2012, who approved obtaining-and-using the samples for this research (SLP/09-2020). Samples from Jalisco came from the Laboratory for the Diagnosis of Emerging and Reemerging Diseases (LaDEER) of the University of Guadalajara, compliant with local ethical authorities. Obtaining-and-using research samples from Jalisco were approved by the *Comités de Investigación, Ética en Investigación y Bioseguridad de la Universidad de Guadalajara*, led by Dr Bárbara Vizmanos Lamonte (CI-00821 and CI-00721). All samples were treated and contained by international standards of Interim Laboratory Biosafety Guidelines for Handling and Processing Specimens Associated with Coronavirus Disease 2019 (COVID-19), and all positive diagnostics were informed to patients and the corresponding Health local authorities between 24 and 48 hours of sampling. An informed written consent for the use of surveillance samples was obtained from all patients.

## RESULTS

### Sampling of asymptomatic SARS-CoV-2 carriers

It was relatively straightforward to have access to samples from Covid19 patients approaching the diagnostics laboratory or the hospital (samples from San Luis Potosi and Jalisco States), but this was not the case for asymptomatic carriers of the SARS-CoV-2 virus from Guanajuato. We solved this problem by tracing back contacts of patients with symptoms and by using a *de novo* self-sampling scheme to collect samples. We developed a highly efficient 3D polycarbonate swab that ensured capture of larger amounts of nucleic acids than what is obtained with traditional cotton swabs (or other materials), greatly reducing the risk of false negatives. The performance of the swab and self-sampling method was validated with 58 volunteers subject to nasopharyngeal sampling by a medical professional and self-sampling following simple instructions provided by us (see Methods and supplementary **Figure S1**).

The self-sampling approach yielded a minimum amount of 20 ng/µl per extraction, carried out in duplicates. We obtained sufficient nucleic acids to perform several RT-qPCRs for separate diagnostic purposes or mutant screening, as well as the entire sequencing of their genomes. By following this approach, we also reduced the risk among members of the potential clusters investigated, and health professionals were under less viral exposure. For the State of Guanajuato, a total of 23 asymptomatic SARS-CoV-2 carriers, plus a total of 37 patients that eventually developed symptoms, could be identified after a screening of 484 individuals. Related metadata and genomic SARS-CoV-2 IDs are provided in Supplementary **Table S2**.

### Targeted RT-qPCR identification of variants of concern in Mexico

We adopted an RT-qPCR approach to identify mutations of concern previously detected in VOCs circulating worldwide, as previously (Vega-Magaña et al. 2021). We screened for the Spike protein mutations E484K, N501Y and 69-70 deletion. A total of 1560 SARS-CoV-2 positive samples from patients coming from Mexico City (60) and the States of San Luis Potosi (170) and Jalisco (1330) from the pre-vaccination stage were characterized. This effort allowed us to identify only one sample during January 2021 (frequency of 0.59%, 1/170) from the State of San Luis Potosi (supplementary **Figure S2-B**). The genome sequence of the associated SARS-CoV-2 virus was obtained by Mexican authorities (Mexico/SLP-InDRE_454/2021, GISAID EPI ISL 1219714), confirming the mutation E484K; but also, independently by us in this study, as the raw reads were required for our population genomics analysis. Likewise, following the same approach, a total of nine independent samples containing the E484K mutant could be detected in samples from the State of Jalisco (supplementary **Figure S2-A**). The genome sequences of three of these samples were generated by Mexican authorities (Mexico/JAL-InDRE_371, 372, 373/2021,GISAID EPI ISL 1093145,1093146,1093147), and us, including the remaining six samples needed for our population genomics analyses (Mexico/JAL-LaDEER-145365, 145340, 139093, E39931, 133706, 147248/2021,GISAID EPI ISL_1360409, 1360408, 1360412, 1360411, 1360407, 1360410). Sample collection of the Jalisco sequences (frequency of 0.69%, 9/1330) occurred during January 2021.

### Phylogenomics reveals convergent evolution and spread of mutations of concern in Mexico: identification of VOI P.4 and B.1.1.222 / B.1.519

To further identify potential mutations of concern and characterize their phylogenetic associations, as well as their co-occurrence and frequency at the clade level, we constructed an *ad hoc* database for Mexican genome sequences, called Mexstrain, which includes selected references to allow for the correct establishment of phylogenetic relationships in the Nextstrain platform. The Mexstrain version used for our analyses consists of 4242 genome sequences (1554 from Mexico), and includes our newly generated high-quality genome sequences from asymptomatic carriers (5) and symptomatic patients (30) from Central Mexico (Guanajuato, San Luis Potosi and Jalisco States). As such, Mexstrain provides the universe of currently available genomic data for the pre-vaccination stage (March 2020 - February 2021) of the Covid19 Pandemia in Mexico, allowing us to identify the presence of additional Spike protein mutations of concern in two sequences or more, at positions 13 (S → I), 18 (L → F), 20 (T → I), 26 (P → S), 144 (144 → X), 152 (W → C), 190 (R → M), 417 (K → N), 452 (L → R), 477 (S → N), 614 (D → G), 677 (Q → H), 681 (P → R/H) and 1176 (V → F). These samples are distributed throughout different clades and/or emerging lineages (**Table 1**).

**Table 1.**
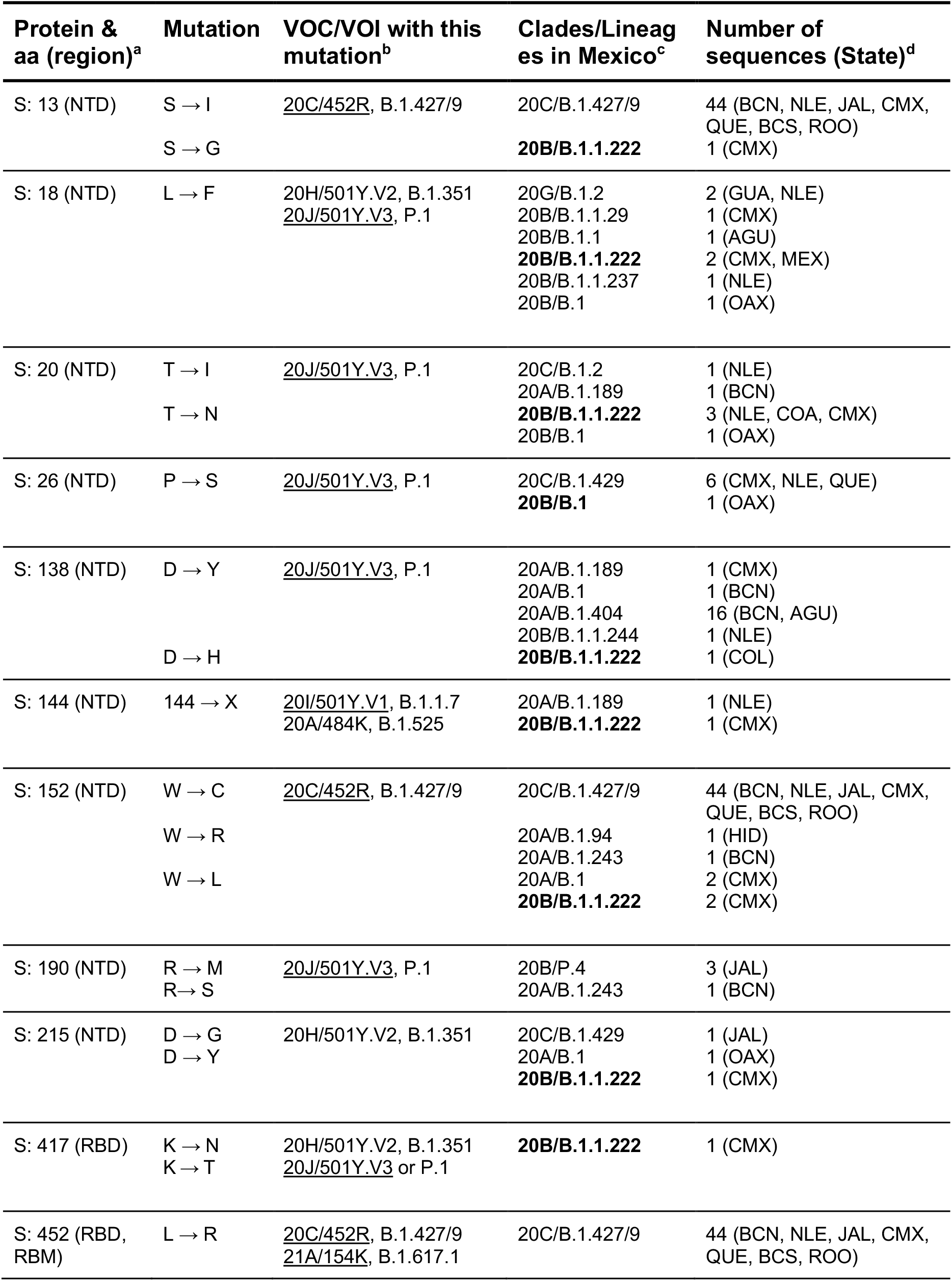
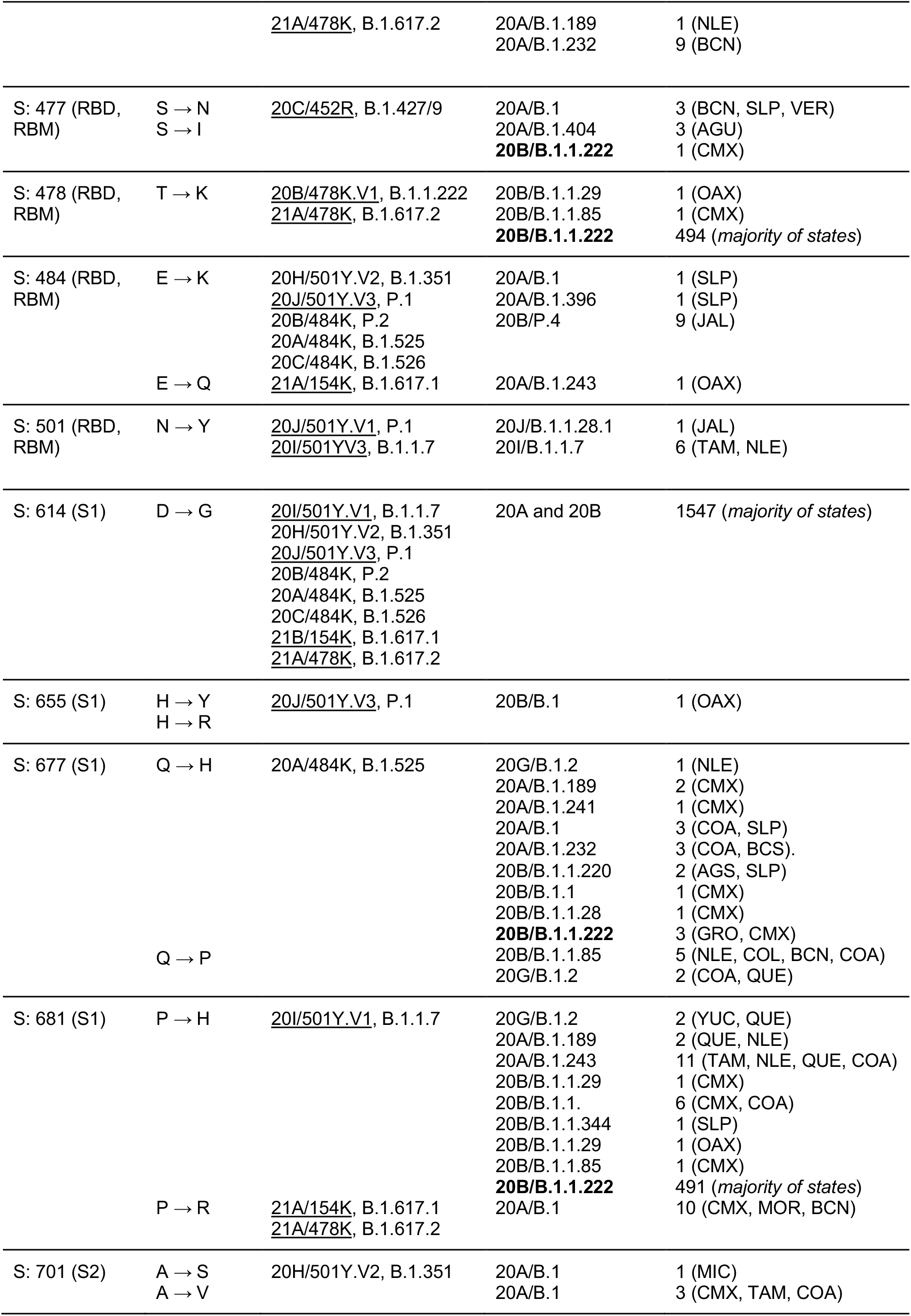
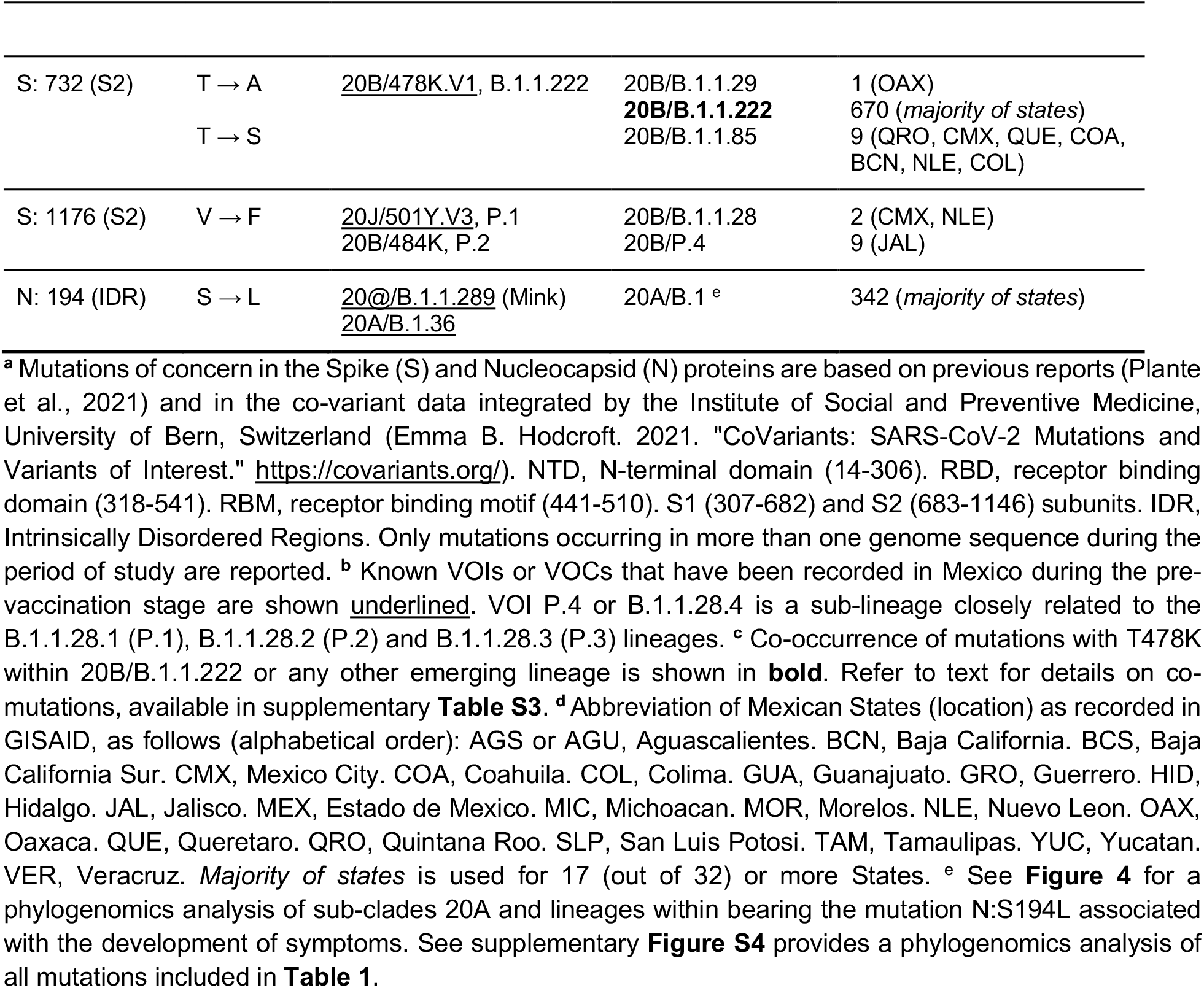
Mutations of concern with multiple occurrences in Mexico during the pre-vaccination stage of the Covid19 pandemia.

More specifically, the genome data generated by Mexican authorities and other laboratories include sequences with the mutation E484K in the context of known VOCs or VOIs. This include (i) one single sequence associated with 20J/501Y.V3 or P.1 (Mexico/JAL-InDRE_245/2021, GISAID EPI ISL 1008714); (ii) as mentioned in previous section, three sequences from Jalisco annotated as 20B/P.2 or B.1.1.28.2 variants (Mexico/JAL-InDRE_371, 372, 373/2021) first identified by us and sequenced in parallel together with further six sequences also identified by us (Mexico/JAL-LaDEER-133706, 139093, 145340, 145365, 147248, E39931/2021; and (iii) two sequences from the State of San Luis Potosí (Mexico/SLP-InDRE_192/2020 and Mexico/SLP-InDRE_454/2021) which belong to Nextstrain clade 20A (different sub-lineages within the Pangolin lineage B.1). This observation make the sequences of scenario (iii) closer to the recently identified E484K-containing lineage 20A/S.484K (B.1.525) (Lasek-Nesselquist et al. 2021) and B.1.243.1 (Skidmore et al. 2021), than to 20B/P.2 (B.1.1.28.2). Both B.1.525 and B.1.243.1 were first reported in the USA, North East States and Arizona, respectively. As discussed in the final subsection of the Results, early cases of lineage B.1.243.1 were present in Mexico during the pre-vaccination stage.

The above-mentioned scenarios (ii) and (iii), together with further E484K-containing variants identified by RT-qPCR screening (supplementary **Figure S2**), places the States of San Luis Potosi and Jalisco as epicenters of E484K-containing SARS-CoV-2 VOI in Mexico. P.1 and P.2 share mutations E484K, D614G and V1176F (the latter appearing to be specific to these closely related emerging lineages), but not N501Y. As expected from independently evolving VOC, mutation V1176F is absent from the two sequences from the State of San Luis Potosi. In contrast, all samples from Jalisco State, scenario (ii), are situated in the same lineage as 20B/P.2 or B.1.1.28.2. The sequence Mexico/JAL-InDRE_373/2021 includes the mutation R190M, an alternative version of mutation R190S present in the VOC P.1 or B.1.1.28.1, which may be signs of evolution in Mexico. Indeed, all Mexican sequences form a distinctive clade supported by the mutations ORF1a: V1071A, P1810L, S3149F (**Figure 1C** & Supplementary **Figure S3**). Based on these observations, and the fact that a similar situation has been reported in The Philippines involving the VOI P.3 (B.1.1.28.3, theta, Tablizo et al. 2021), we propose to re-name the Jalisco P.2-like variant as P.4 or B.1.1.28.4. Given that P.3 has accumulated six further dangerous mutations in the Spike protein, including E484K, N501Y and P681H, further investigation of P.4 in Mexico due to its evolutionary potential is recommended.

**Figure 1.**
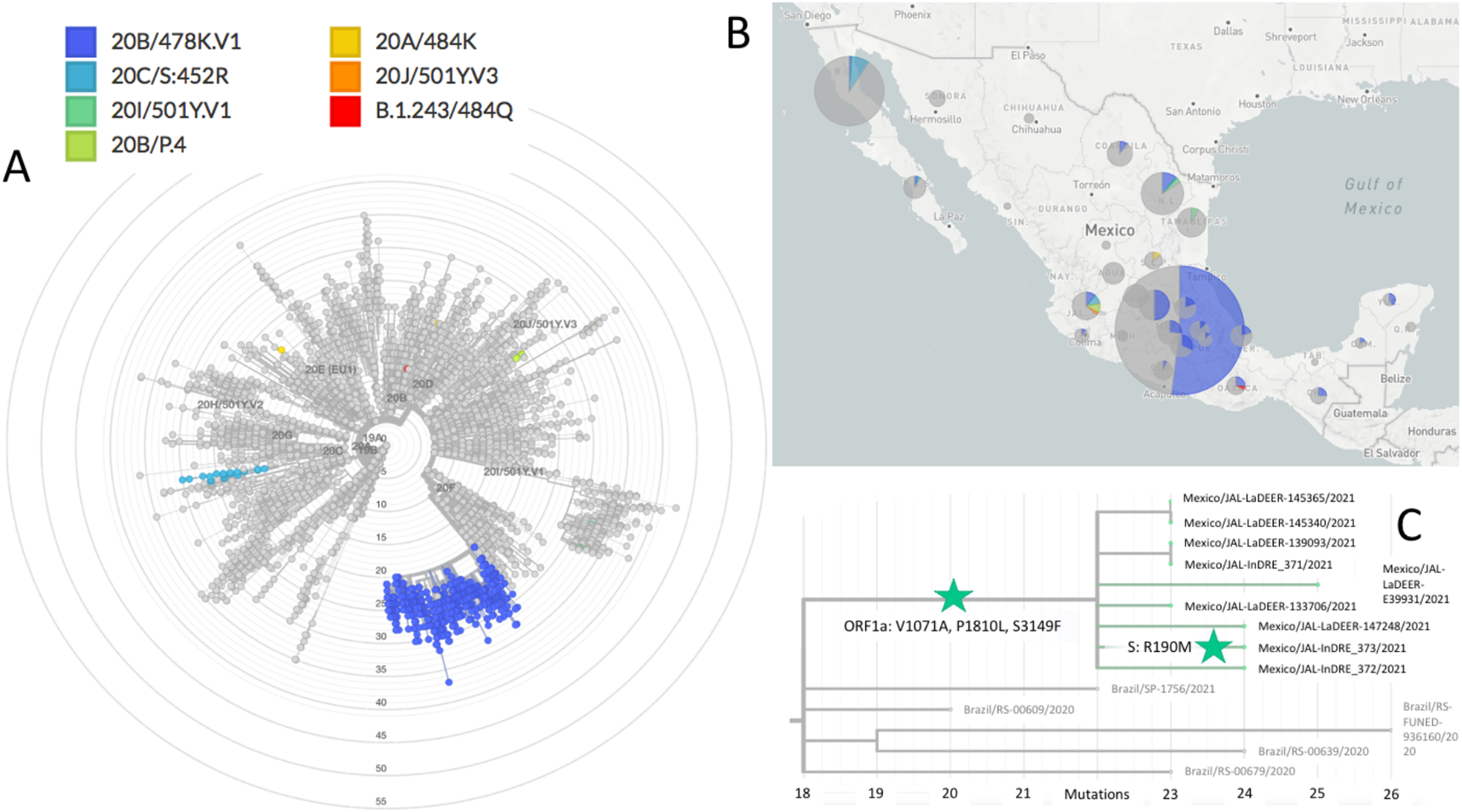
Phylogenomics of SARS-CoV-2 variants during the pre-vaccination stage in Mexico. **A**) The Mexstrain phylogenomic tree showing VOI/VOC identified during the pre-vaccination stage. 20A/484K refers to the San Luis Potosi samples, belonging to lineages B.1 and B.1.319. **B**) Geographic distribution of VOI/VOC in Mexico. **C**) 20B/P.4 or 20B/B.1.1.28.4 evolving in Jalisco, Mexico, from P.2. ORF1a and Spike protein amino acid mutations (ORF1a: V1071A, P1810L, S3149F; S: R190M) are indicated with green stars.

Our phylogenomics analyses also revealed a different 484 mutation involving another amino acid substitution, namely, E484Q in the sequence Mexico/OAX-InDRE-61/2020 (August 2020, GISAID EPI ISL 576264). This sequence in shown in **Figure 1A** and it corresponds to lineage B.1.243, further discussed below in the population genomics subsection. Glutamine (Q) is a basic residue, and therefore it is tempting to speculate that a similar effect to that generated by the basic residue lysine (K) may be in place, as proposed for the recently designated VOI 21A/S:154K evolving in India (B.1.617.1, kappa, Ferreira et al. 2021). Irrespective of the codon usage of SARS-CoV-2 (Sheikh et al. 2020), the mutations from E to K (G → A) and from E to Q (G → C) involve single and non-sequential nucleotide point mutations, and thus they may offer alternative solutions to the same phenotype. A similar scenario with multiple combinations at the codon level, within different lineages in Mexico, can be seen in the mutation of concern S477N/I (Singh et al. 2021). These mutations happen in seven sequences from different States distributed throughout different lineages (B.1.404 and B.1, for instance), and they involve mutations G → A and G → U in the second letter, respectively, which can be generated from two different sets of codons. The case of mutation 681 from P to R/H, present in the Mexican VOI 20B/478K.V1 discussed next, also provides an important example of the SARS-CoV-2 genomic plasticity, concomitant with convergent evolution of mutations in the Spike protein (Cherian et al. 2021). These cases, and others included in **Table 1** and supplementary **Figure S4**, raise the importance of the characterization of allelic variation *de novo* as we do in the final population genomics subsection.

Mutagenesis of concern does not occur solely within the RBD of the Spike glycoprotein, but also in its N-terminal domain (Suryadevara et al. 2021), which is rich in glycosylation sites (Watanabe et al. 2020). For instance, mutation L18F is a mutation of concern as it happens in variants 20H/501Y.V2 or B.1.351 and 20J/501Y.V3 or P.1 and has been implicated in antibody scape. Within the period of analysis, this mutation occurs in Mexico in two forms: (i) in the single case associated with 20J/501Y.V3 or P.1 previously mentioned; and (ii) in two different lineages within several clades, accounting for at least six different backgrounds in a total of eight samples. A similar scenario for the NTD deletion 144X can be found in four samples from the North East of Mexico, Nuevo Leon and Tamaulipas States, associated with 20I/501Y.V1 or B.1.1.7, and more importantly, in seven samples identified in two different lineages, 20A/B.1.189 and 20B/B.1.1.222. These examples of convergent evolution of mutations of concern, together with many others, are also included in **Table 1** and supplementary **Figure S4**.

In agreement with public announcements of Mexican Health authorities, a genomic lineage belonging to 20B/B.1.1.222, eventually leading into 20B/B.1.1.519, was confirmed simultaneously to the cognate report (Rodríguez-Maldonado et al. 2021). Worryingly, these lineages have in common between them, and with the recently designated VOC 21A/S:478K (B.1.617.2, delta, Ferreira et al. 2021), first identified in India and shown to undergo immune escape, mutations T478K and P681H/R. In addition, the Mexican variant shows mutation T732A. More than one third of available sequences with a nationwide distribution with these mutations could be identified (**Figure 1 & 2** and supplementary **Figure S4**). Although occurrence in Mexico City seems higher, the earliest sequence with these three mutations (annotated as lineage B.1.1.29) is from the South East State of Oaxaca, recorded as early as July 2020 (Mexico/OAX-InDRE_258/2020, GISAID EPI ISL 1054990).

**Figure 2.**
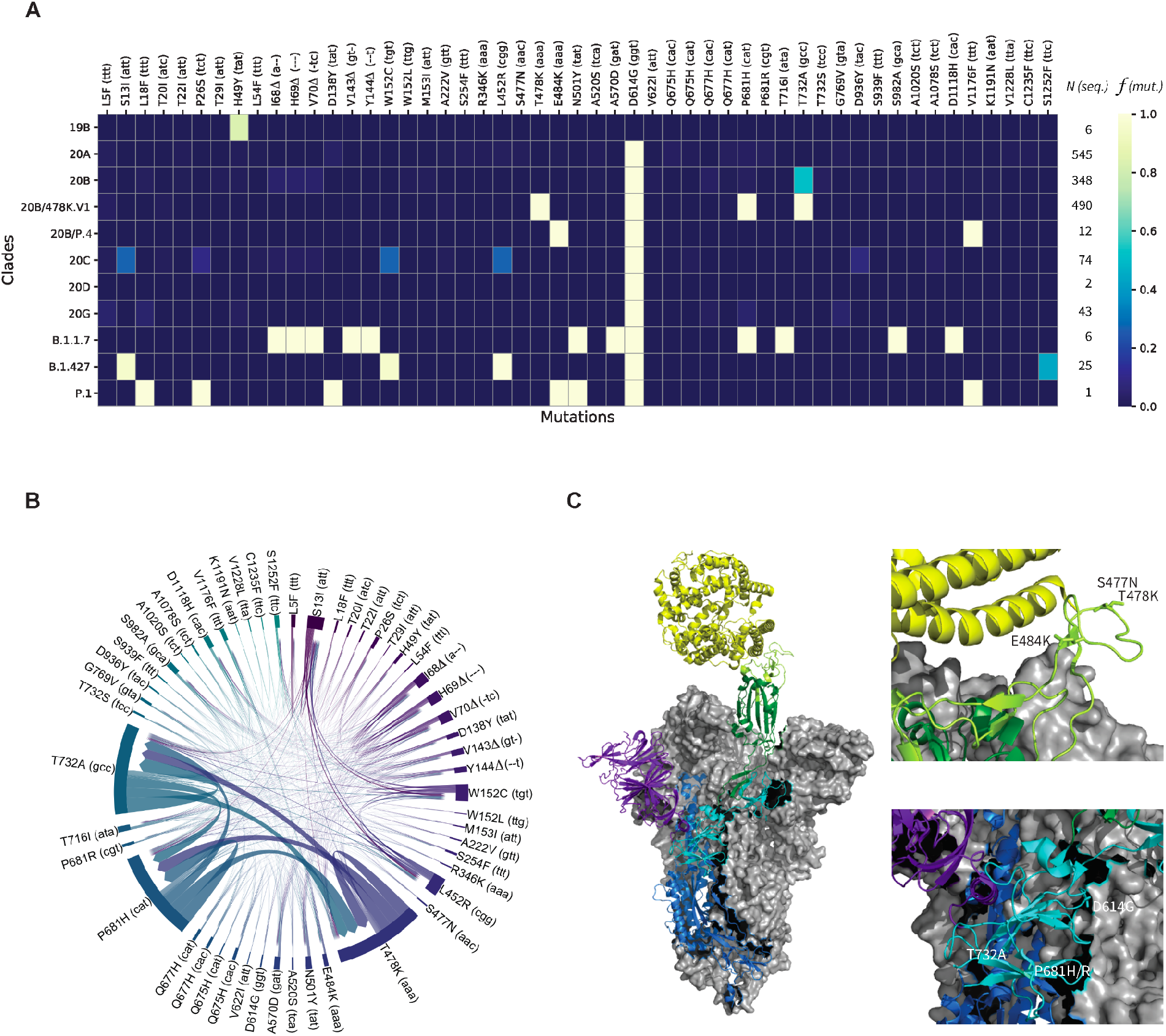
Systematic sequence/structure analysis of Spike protein mutations of concerns. **A**) The heatmap highlights relative frequencies *f* for 51 important mutations (out of 315) across 1552 viral genomes arranged in 11 clades (19B, 20A-D, 20B/478K.V1, 20B/P.4, 20G, B.1.1.7, B.1.427, P.1). *N* indicates the number of available genome sequences per clade. See supplementary **Figure S5** for a heatmap including all mutations. **B**) The chord diagram displays the covariances between the 51 most frequent mutations. The thickness of the arrows denotes the strength of a covariance. **C**) *Left* - Electron Microscopy structure (PDB:7A94 ref) of the trimeric Spike (S) glycoprotein of SARS-CoV-2 bound to one Angiotensin-Converting Enzyme-2 (ACE2, yellow). Two chains of the viral protein are shown in gray surfaces. The third chain comprises the N-terminal domain (purple), the S1 (cyan) and S2 (blue) protein subdomains, the Receptor-Binding Domain (RBD, green), and the Receptor-Binding Motif (RBM, light green). *Right* - Positions of the important mutations S477N, T478K, E484K, D614G, P681H/R, and T732A in two different regions.

We then systematically investigated overall mutation’s frequency and co-occurrence in lineages B.1.1.222 and B.1.1.519. For this, we first identified 315 mutations of concern in the Spike protein that occur in at least one genome sequence during the period of analysis. We then analyzed their incidences and their covariances across the 1552 genome sequences in which these mutations occur. The results of this analysis are presented in **Figures 2A & B** and supplementary **Figure S5. Figure 2A** highlights the incidences of the 51 most frequent mutations of concern within the 11 clades used to organize this data. We then selected the 51 mutations that were identified in at least five genome sequences overall, indicating the number of available genome sequences (*N*) per clade and the incidence of each mutation expressed as its relative frequency (*f*), also per clade. The highest incidences are depicted in yellow (close to 1.0), while low relative frequencies or inexistent mutations are shown in shades of blue.

These analyses show that roughly 80% of the clade 19B sequences display the Spike protein mutation H49Y, which appears to be unique to that clade. The insertions or deletions of the other mutations of concern illustrate the evolutionary relationships between subsequent clades, starting with clade 20A. Besides the conserved D614G mutation, sequences from the newly detected and proposed VOI 20B/478K.V1 and 20B/P.4, share, in different combinations, mutations T478K, E484K, P681H/R with VOC alpha, gamma and delta (20I/B.1.1.7, 20J/P.1 and 21A/B.1.617.2). Variant 20B/478K.V1 has in addition the mutation T732A. **Figure 2B** further validates the co-occurring mutations of concern mentioned above, particularly between T478K, P681H, and T732A, from the dominant clade 20B of variant 20B/478K.V1. With only 25 sequences available, this analysis also allowed noting for weaker covariances between S13I, L452R, and W152C of the VOI B.1.427/9.

Among these results, although not at a high frequency, cases of T478K co-occurring with other Spike protein mutations of concern could be detected, including: (i) K417N, which is present in 20H/501Y.V2 or B.1.351, identified in one sample (February 2021, GISAID EPI ISL 1137473); (ii) L18F, which is present in 20J/501Y.V3 or P.1, identified in one sequence from the State of Oaxaca, in the parental lineage B.1 (January 2021, GISAID EPI ISL 1168605); and (iii) the deletion of amino acid in position 144, which occurs in 20I/501Y.V1 or B.1.1.7 and 20A/S.484K or B.1.525, in one sample from Mexico City (February 2020, GISAID EPI ISL 1181713). Similar to convergent evolution of E484K leading to the potential VOI from the State of San Luis Potosi, mutation T478K is not only present in the entire country, but the sequence Mexico/OAX-InDRE_535/2021 (January 2021, GISAID EPI ISL 1168605) has it, and belongs to 20B/B.1. It is also interesting to note that mutations T478I/R, previously identified as dangerous (Li et al. 2020; Muecksch et al. 2021), could not be found in Mexico during the period of analysis.

### 3D structural analysis of dangerous mutations occuring in Mexican VOI P.4 and B.1.1.222 / B.1.1.519

To predict the alleged roles the detected mutations of concern may take in the transmission and/or viral replication, we localized their positions onto the glycoprotein Spike of SARS-CoV-2. For this, we focused on S477N, E484K and D614G as previously identified mutations of concern; and in mutations T478K, P681H/R and T732A present in the proposed VOI 478K.V1. We used the electron microscopy trimeric structure (PDB:7A94) published by Benton and co-workers (Benton et al. 2020). These authors reported several structures free or bound to one or several Angiotensin-Converting Enzyme-2 (ACE2), of which we picked the example bound to a single ACE2 for simplicity, as shown in **Figure 2C**. In that figure, two of the three chains within the glycoprotein S are depicted in gray structures to keep our attention to the single chain interacting with ACE2 (yellow). The third chain includes the N-terminal domain (purple), the S1 (cyan) and S2 (blue) protein subdomains, the Receptor-Binding Domain (RBD, green), and the Receptor-Binding Motif (RBM, light green). Three mutations of concern S477N, T478K, and E484K, are in direct vicinity with the human receptor ACE2, located within the RBM. The mutations D614G and P681H/R are found in the S1 protein subdomain near T732A, taking part in S2. Overall, these structural projections emphasize the expected risks associated with these mutations, in particular for the proposed VOI 478K.V1 emerging in Mexico.

### Population genomics of SARS-CoV-2 in Central Mexico leads to the discovery of the potential VOI B.1.243

Our reference-based (NC_045512.2) approach yielded high-quality read alignments for 85 of the 102 sequence libraries with a median value of uniquely mapped reads of 177,688 and 1st and 3rd quartiles of 16,292 and 1,751,201, respectively (supplementary **Table S3**). These alignments were distributed in 44 libraries from Guanajuato, 32 from San Luis Potosí and 9 from Jalisco. Once polymorphic sites were filtered by sequencing depth, quality of the alignments and population frequency, 82 (97%) of the 85 initial samples were retained. From a total of 500 polymorphic sites across these samples, 59 high-quality SNVs/indels were retained, spanning the ∼30 Kb of the SARS-CoV-2 reference genome. These polymorphic sites include 53 SNV and six indels ranging from two up to seven nucleotides in length. This result shows that the use of raw reads from multiple genomes increased the predictive power of genomic variation, in particular for low-frequency variants that could be overseen in lower sample size experiments (Wang et al. 2021).

Our samples contained SNVs in one of the early strain types (VI) that spread out of China (C241T, C3037T, C14408T and A23403G mutations) which has the haplotype of allelic associations 241T-3037T-14408T-23403G, detected in high frequency worldwide (December 2020, Korber et al. 2020) and that can be involved in increasing the fitness of the SARS-CoV-2 virus (Yang et al. 2020). Overall, we found that the majority of the genomic variants (SNVs + indels) occur in low frequencies, with the exception of a few point variants, such as P323L in the RdRp protein; E484K, D614G and V1176F spanning the Spike protein region, and R203K and S194L in the Nucleocapsid region (**Figure 3A & 3B**). Importantly, we found the mutation T478K in 4 of the 31 San Luis Potosi genomes, but it was filtered out of the population genetic analyses due to its low population frequency (< 5%). The mismatch distribution of the per-polymorphic site population frequencies (**Figure 3C**) suggests that there is selection on multiple mutations that result in several high frequency variants in the population.

**Figure 3.**
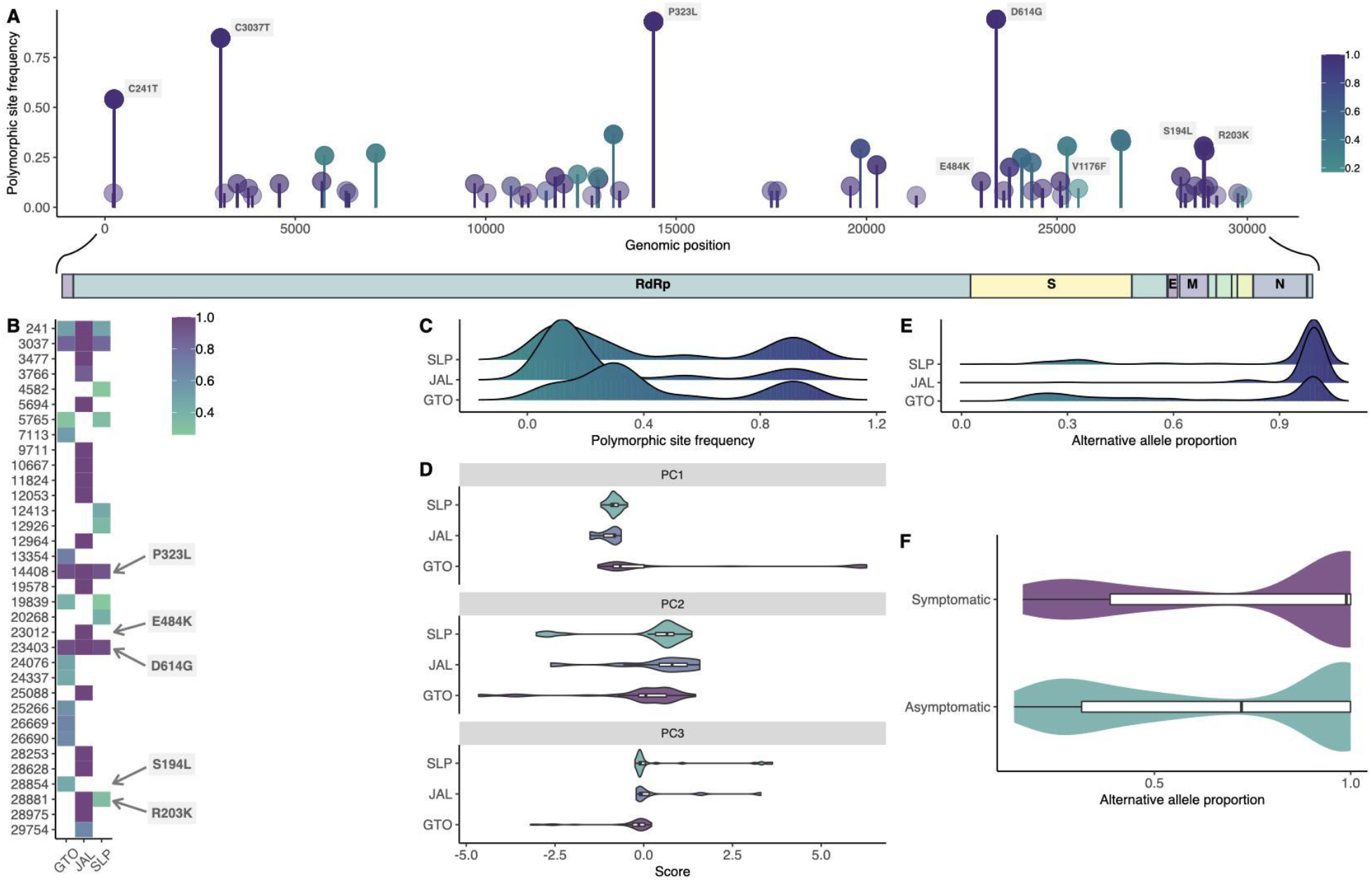
Landscape of genomic variation unveiling the fixation of mutations in central Mexico. **A**) The genome-wide population frequency of SNVs is plotted as lollipops. Darker colors reflect the per-site median allelic imbalance. **B**) Heatmap showing the relative allele frequencies of the most common variants in the population in a per-locality fashion. **C**) Density plots showing the distributions of the allele frequencies of the 59 high-quality polymorphic sites for each locality. **D**) The three principal components of genetic covariance among samples are shown as violin plots for each location, suggesting that samples from Guanajuato (GTO) have a genetically distinct composition from those of Jalisco (JAL) and San Luis Potosi (SLP). **E**) The distributions of the per-variant intra-host allelic imbalance are shown as density plots for each locality. **F**) The violin plots depict the distributions of allelic imbalance for symptomatic and asymptomatic hosts within Guanajuato.

To evaluate the extent of substructure in our sampling, we used the 82 samples with high-quality polymorphic sites to generate a genetic covariance matrix representing the relatedness among samples, which in turn was used to perform a PCA. The principal components PC1, PC2 and PC3 explained the 52%, 21% and 15% of variance genetic differences, respectively. The PC1 suggests overall admixture among the three localities with the exception of a small portion of the samples from Guanajuato separated from those from San Luis Potosi and Jalisco (**Figure 3D**) despite the presence of different lineages and variants. By evaluating the allelic imbalance, as a quantitative measurement of the co-occurrence of variants within hosts we found that for the majority of polymorphic sites, the derived allele is found in higher proportions (**Figure 3E**), mostly in Jalisco, likely explained by the pre-screening of the E484K in this locality. In some polymorphic sites that were extracted from non-screened genomes, namely in Guanajuato, the ancestral allele dominates the sequencing reads in the sample.

The Guanajuato samples include asymptomatic patients, which could help explain the smaller cluster that we found (**Figure 3D & 3E**). To further investigate the role of viral genetic composition in the presence or absence of symptoms in the Guanajuato cluster, we compared the median values of the allelic imbalance between symptomatic patients and asymptomatic carriers. We found that symptomatic patients have higher proportions of the derived allele (Wilcoxon test; Effect size = 0.12, *T* = 17,808, p = 0.00692) (**Figure 3F**). By contrasting the allelic imbalance between symptomatic and asymptomatic hosts in a polymorphic site fashion, we only found a significant association of the C28854T mutation (ANOVA; *F* = 20.5; p = 0.003) (**Figure 3A**). This SNV corresponds to the amino acid change S194L in the Nucleocapsid protein and has a population frequency of 30% in Guanajuato.

Motivated by these results, we analyzed the prevalence of mutation N:S194L in the extended Nextstrain phylogeny, and found three major sub-clades within the 20A clade with high prevalence: (i) two closely related sub-clades dominated by the B.1 lineage, where most of the Mexican genomes are placed, including more than one fifth of all sequences available for the period of analysis (342 out of 1554); and (ii) a mainly Asian sub-clade that includes the emerging lineage B.1.36.# (where # refers to many sub-lineages, e.g. 6, 10, 16, 18, 19, 21, 22, 25, 26 and 27) (**Figure 4A**). However, many different Pangolin lineages are annotated within the implicated N:S194L-containing 20A sub-clade of scenario (i) (**Figure 4B**). In addition to the B.1 lineage within these two 20A sub-clades, lineage B.1.243, with 78 genome sequences sharing the point mutation N:S194L and the deletion NS9c:Q41* (22.8% of total sequences within these two 20A clades and 5.1% of the total genome sequences), caught our attention. Analogous to the E484K mutation present in the parental lineage B.1.36 of scenario (ii), which has been implicated in regional outbreaks in India (Pattabiraman et al. 2021), the proposed new Mexican lineage of scenario (i) includes the two San Luis Potosi E484K-containing variants identified after RT-qPCR screening (lineages B.1 and B.1.319), and the Oaxaca E484Q variant appearing early on during the pandemia, which corresponds to lineage B.1.243. The potential for evolving the dangerous E484K/Q mutation within this Mexican lineage is supported by the recent detection of the sub-lineage B.1.243.1 in Arizona, USA, which includes mutation E484K (Skidmore et al. 2021). All together, these observations have led us to propose the lineage B.1.243 in Mexico as a VOI, which we believe warrants further investigation and direct surveillance as it accumulates dangerous Nucleocapsid *and* Spike protein mutations during the vaccination stage.

**Figure 4.**
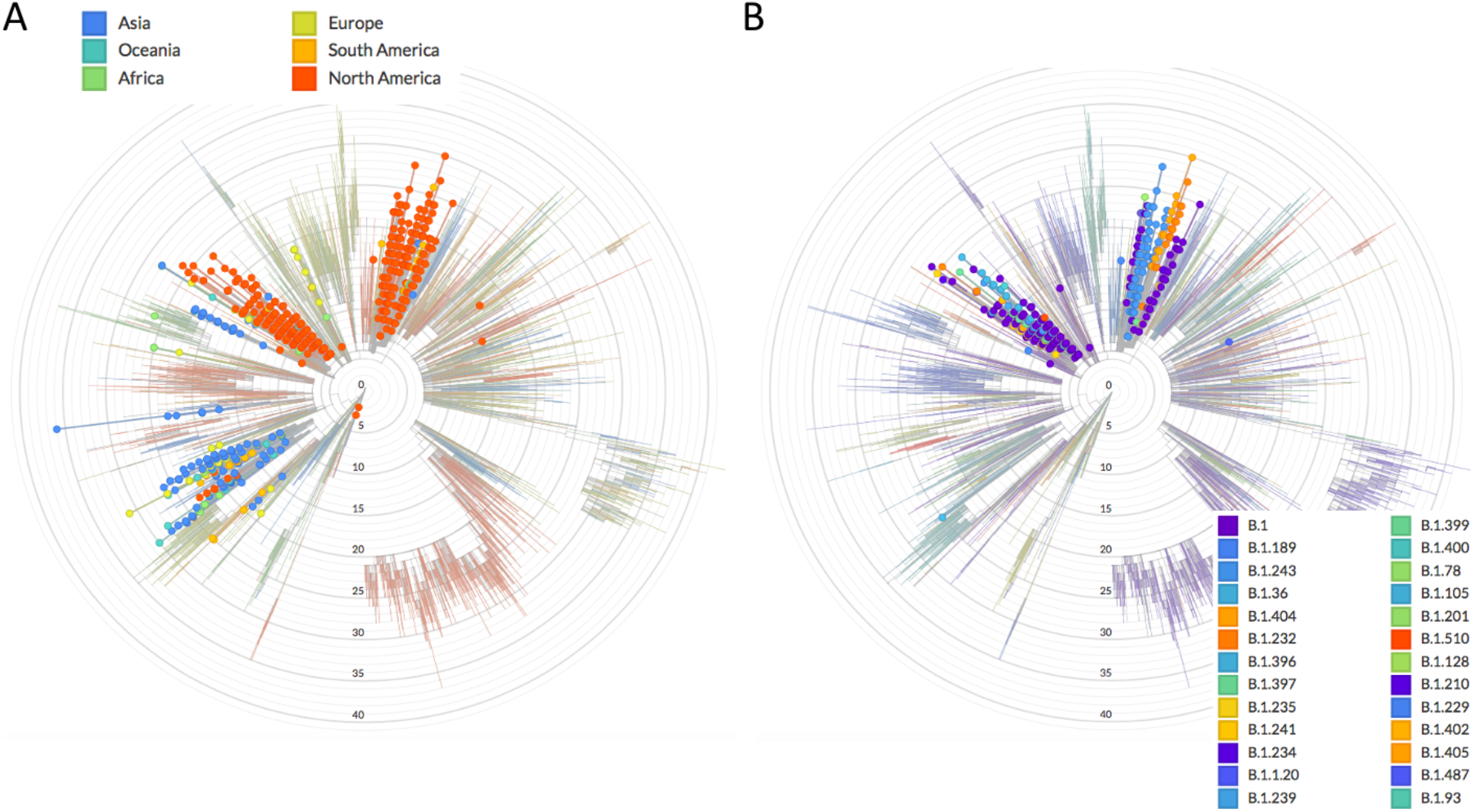
Phylogenomics of N:S194L associated with symptomatic versus asymptomatic carriers. **A**) Worldwide occurrence of N:S194L mutation using NextStrain **B**) Same analysis highlighting only sequences from Mexico distributed in two sub-clades consisting of a broad diversity of Pangolin lineages, dominated by B.1 but also showing B.1.243 with the potential to evolve the E484K mutation.

## DISCUSSION

At the moment of emergence of all SARS-CoV-2 variants a massive or global immune pressure elicited by vaccination or antiviral treatments was not present. Indeed, it has been reported that reinfections of SARS-CoV-2 can occur (Tomassini et al. 2021; Zhang et al. 2020; Kellam & Barclay 2020; Lu et al. 2020). Rapid and independent viral evolution has been observed in other pathogens such as the respiratory syncytial virus (RSV). In 2009, during the influenza A/H1N1 pandemic, a 72-nucleotide duplication in the G gene of the RSV-A occurred. During the following three years multiple independent duplication events occurred around the world (Comas-García et al., 2018). In fact, convergent evolution of the gene G in RSV-A and RSV-B have been reported (Muñoz-Escalante et al. 2019). The mechanism that drives RSV and SARS-CoV-2 convergent evolution may be similar. This may involve the lack of a specific antiviral treatment, the lack of vaccines (SARS-CoV-2 convergent events occurred before the distribution of vaccines), a similar R0 of both pathogens, a global high transmission rate and the capacity to escape from the humoral immune response.

During the pre-vaccination stage in Mexico, our analyses revealed two potential VOI with mutations in the Spike protein: 20B/478K.V1 or B.1.1.222 / B.1.1.519 and P.4 or B.1.1.28.4, plus an evolutionary event with the potential to lead into a B.1.243 sub-lineage leading to a third VOI. On one hand, although further data is needed to confirm the E484K-containing sequences detected in San Luis Potosi as VOI, which occurs within a clade dominated by the B.1.243 lineage, our results emphasise the potential of a combined targeted RT-qPCR screening and genome sequencing approach to anticipate epidemiological hotspots. It is tempting to speculate that these sequences could be related to the origin of the E484K-containing variant B.1.243.1 detected in Arizona (Skidmore et al. 2021), a USA Southern state that borders the Mexican Northern States of Sonora and Chihuahua. However, further data is required to assess the phylodynamics of this lineage and distinguish between common origin or convergent evolution. In the meantime, it is clear that the San Luis Potosi E484K-containing variants are different to the closely related and previously reported VOC/VOI P.1, P.2, and recently, P.3 sequences (Tablizo et al. 2021). Although the proposed P.4 variant still appeared at a low frequency (0.68%) during the period of analysis, it can be concluded that this variant is the result of an early P.2 entry followed by local evolution (including mutation of R190 into M instead of S). These results are in line with data suggesting convergent evolution and global spread of the virus containing the E484K mutation during the pre-vaccination stage (Lasek-Nesselquist, Lapierre, et al. 2021; Annavajhala et al. 2021; Ferrareze et al. 2021; Cherian et al. 2021).

On the other hand, 20B/478K.V1, with mutations T478K, P681R/H and T732A in its spike protein, can undoubtedly be classified as a VOI, with the potential to become a VOC. This has been suggested as well by other authors (Di Giacomo et al. 2021; Rodríguez-Maldonado et al. 2021; Lasek-Nesselquist, Pata, et al. 2021), who simultaneously reported the occurrence of this variant in Mexico, USA and Europe. Importantly, during the pre-vaccination stage, this VOI was ascribed as the B.1.1.222 Mexico and USA lineage, which at time of submission appeared more often as B.1.1.519. Beyond nomenclature, the phenotypic implications of this VOI, including potential clinical outcomes and impact upon diagnostics, represent a pressing issue that remains to be addressed. Of particular concern are the worrisome Covid19 circumstances in India, which may relate to the recently demonstrated ability of the rapidly emerging VOC 21A/B.1.617.2 to escape the immune system (Ferreira et al. 2021). Similar to variant 20A/478K.V1, the latter VOC includes the mutations T478K and P681H/R. Indeed, our 3D structural analysis of the spike protein hints towards two regional hotspots for viral mutagenesis. The first one is in the flexible loop of the RBD, and includes S477N, T478K, and E484K. These mutations may affect the interactions between the spike protein and the ACE2 receptor (Singh et al. 2021; Muecksch et al. 2021). The other hotspot is within S1-S2 subdomains, which includes the furin-like protease cleavage site (Xia et al. 2020; Starr et al. 2020), and contains D614G, P681H/R and T732A. Mutations T478K and S477N, involving changes from similar hydroxylated side-chains (T and S) into positively charged basic amino acids (K and N) may have analogous functional roles reinforcing the ability of the virus to bind to the human ACE2 receptor (Starr et al. 2020). Interestingly, S477 has been identified as the most flexible amino acid within the RBM and with the largest number of mutations (Singh et al. 2021), which might be stabilized by mutation T478K. A case exemplifying a similar scenario in which physically close mutations contribute towards an analogous structural solution is provided by the Spike protein mutation Q613H. In this case, mutation Q613H defining the emerging lineage A.23.1, from Uganda, Africa, may have taken the role of the mutation D614G in B lineages during evolution of A lineages (Bugembe et al. 2021).

Our population-level results are consistent with previous reports that highlight the dual role of Spike and Nucleocapsid proteins in adaptive evolution of SARS-CoV-2 to their hosts (Wang et al. 2021). Our finding that symptomatic patients associated with the N:S194L mutation is particularly intriguing. The SARS-CoV-2 nucleocapsid protein N is a multidomain RNA-binding protein that is found inside the viral envelope and it is required by the virus to maintain its structure and viability once the virus has entered the cell (Chang et al. 2014; Tomaszewski et al. 2020). The mutation we identified, N:S194L, is within the central domain composed of Intrinsically Disordered Regions (IDRs) (Chang et al. 2014), which are conformationally flexible and promiscuous (Barik 2020), and are increasingly recognized as important in increased viral transmission (Tomaszewski et al. 2020; Forsythe et al. 2021). Mutations in positions 193, 197, 203 and 204 of the linker IDR have been mapped to the loops connecting disordered and structured regions (Tomaszewski et al. 2020), implicating these sites in increased transmission. Interestingly, variants of lineage B.1.36.# with mutation N:S194L have been associated with higher mortality in Gujarat, India (Joshi et al. 2021). Joshi et al found in March 2021 that this variant had an allele frequency of 47.62 and 7.25% in deceased patients, compared to 35.16 and 3.20% of the same variant in recovered patients, from the Gujarat and global datasets, respectively (although the difference was only significant in the global dataset). Additionally, the N:S194L mutation also occurs at a very high frequency (89.9%, N=1270) in lineage B.1.1.289 (Plante et al., 2021), isolated in Denmark and shown to co-infect humans and minks (Oude Munnink et al. 2021). The evolutionary and functional relationships between the Mexican sub-clades bearing this mutation, including lineage B.1.243 (but also other lineages, e.g. B.1, B.1.189 etc.), and the previously reported and partially characterized lineages B.1.36.# and B.1.1.289, remain to be investigated.

A clear advantage of the population-level approach adopted here, is that it allowed us to identify and characterize SNVs from fragmented genomes. This calls for an additional global repository that can be used for population-level studies. As was previously proposed by Yang et al (2020), SNVs may become an important consideration in SARS-CoV-2 classification, surveillance and tracking as viruses travel through the world and accumulate additional mutations by convergent evolution. Our finding of the mutation N:S194L contributes to an additional site for RT-qPCR screening of samples outside of the Spike region. Future population genomic and phylogenomic studies of SARS-CoV-2 must consider the protein multidimensional landscape of the virus, both Spike and Nucleocapsid. Understanding the role of intra-host selection pressures and their subsequent transmission as vaccination continues is critical. For instance, we do not know if new variants occur mostly as the result of admixture, or the mixtures of different variants within the same person, nor their relationship to various vaccines or mixed vaccination in a diverse human background. As different vaccines are used within and across regions, and varying timing of vaccination programs across countries takes place, genome surveillance will still be of essence to monitor and identify emerging phylogenetic lineages that could become VOCs. Foremost, the recent appearance of various sub-strains that result from prolonged individual infections and subsequent transmission, as well as regional surges and episodic outbreaks (Wang et al. 2021) prompt more studies of phylogenomics coupled with population level analyses that can help provide detailed recommendations to decision-makers.

## Data Availability

All data is available in public databases.

http://www.ira.cinvestav.mx/ncov.evol.mex.aspx

https://github.com/luisdelaye/Mexstrain

https://github.com/plissonf/Phylogenomics_SARS-CoV-2_Mexico

## ACKNOWLEDGEMENTS

We would like to acknowledge colleagues and members of the public who provided samples, as well as all authors who make SARS-CoV-2 sequence data available via the GISAID initiative. We are in debt for technical support, access to diagnostics services and/or critical discussions with Francisco de Alba, Carlos Angulo, Guillermo Corona, Eloisa Díaz-Francés, Fernando Fontove, Rafael Franco, Ámbar Gómez, Bruno Gómez-Gil, Ileana Gutiérrez, Gabriel Hernández, Luis Hernández, Alfredo Herrera-Estrella, Silvia Jerez, Beatriz Jiménez, Alejandro Ledesma, Enrique López, Miguel Nakamura, Arturo Nieto, Mizraim Olivares, Hilda Ramos, Victor Rivero, Nelly Selem, Xavier Soberón and Dulce Valdivia. We also thank Ricardo Grande, Francisco Pulido and Gloria Vazquez from the Unidad de Secuenciación Masiva y Bioinformática of the “Laboratorio Nacional de Apoyo Tecnológico a las Ciencias Genómicas (CONACyT no. 260481) for their support in sequencing services. This work was funded by Conacyt, grant No. 313075 to ACJ, and the resources of GrupoT (OGG). FBG is recipient of a Newton Advanced Fellowship, Royal Society, UK (NAF\R2\180631). FP is supported by a Cátedras CONACYT fellowship. Work in the ACG laboratory at UASLP was funded by the income generated by the Clinical Laboratory Diagnostic Unit.

## DATA SUMMARY

The Mexstrain-Nextstrain data used for this study can be found at: http://www.ira.cinvestav.mx/ncov.evol.mex.aspx.

Data generated and used in this study is provided as supplementary Tables:

**Table S1**. Self-sampling performance validation data generated in 58 volunteers.

**Table S2**. Metadata of SARS-CoV-2 genomes generated and investigated.

**Table S3**. Complete dataset of the population genomics analysis of SARS-CoV-2.

