## Supplementary Figures for "Phylogenomics and population genomics of SARS-CoV-2 in Mexico during the pre-vaccination stage reveals variants of interest B.1.1.28.4, B.1.1.222 or B.1.1.519 and B.1.243 with mutations in the Spike protein and the Nucleocapsid"

### Table of Content

**Figure S1.** Self-sampling for collection of samples from asymptomatic SARS-CoV-2 carriers. *Page 2*

**Figure S2.** RT-qPCR screening of E484K mutation in San Luis Potosi and Jalisco. *Page 3*

**Figure S3.** P.3 distinctive sub-clade supported by specific mutations (ORF1a and Spike). *Page 4*

**Figure S4.** Comprehensive Nextstrain analysis of mutations included in **Table 1**. *Pages 5-29*

**Figure S5.** Heatmap of relative frequencies (*f*) of Spike protein mutations in Mexico. *Page 30*

**Table S1.** Self-sampling performance validation data generated in 58 volunteers. *Separate file*

**Table S2.** Metadata of SARS-CoV-2 genomes (82) generated and investigated. *Separate file*

**Table S3.** Complete dataset of the population genomics analysis of SARS-CoV-2. *Separate file*

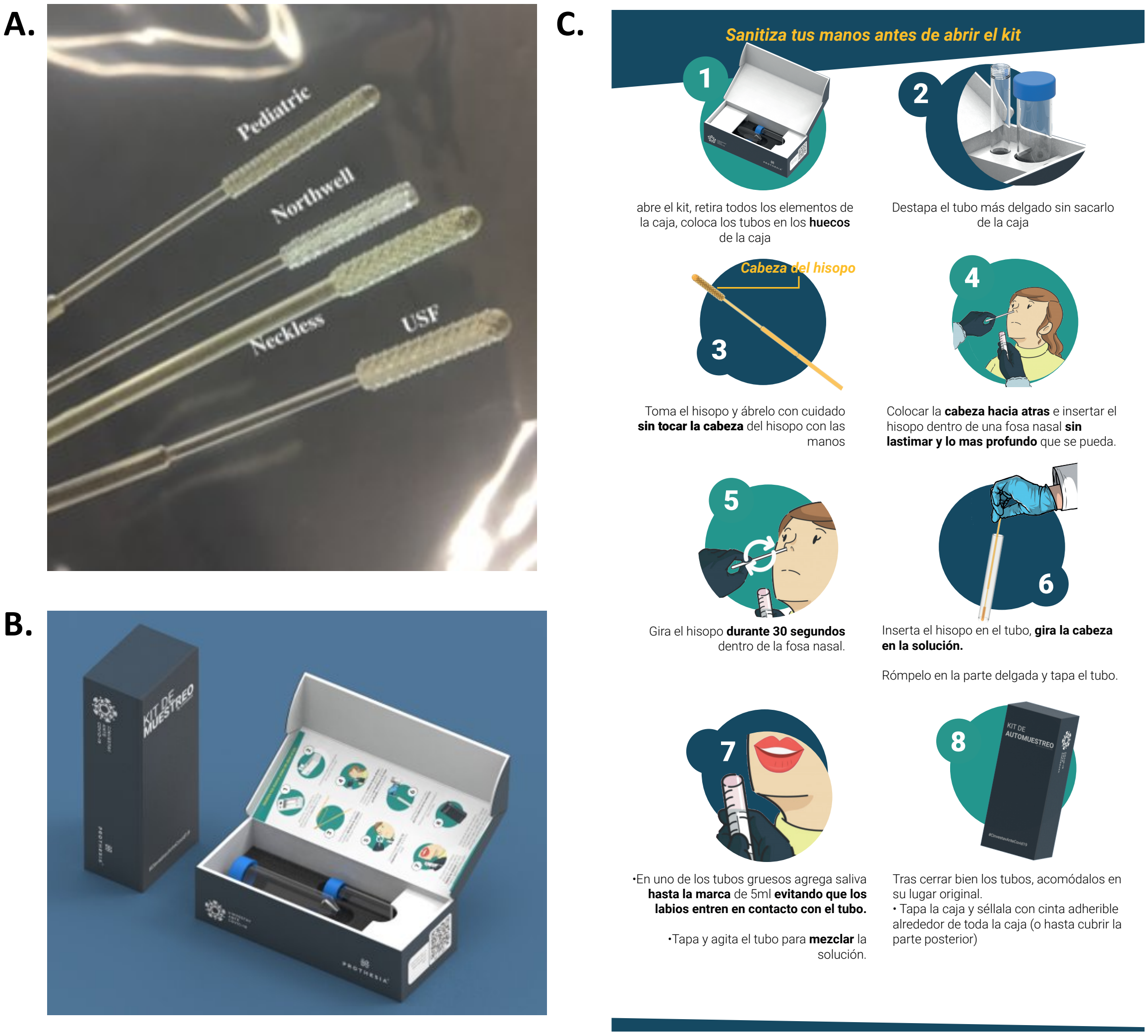


**Figure S1. Self-sampling for collection of samples from asymptomatic SARS-CoV-2 carriers.** **A.** 3D swabs used in this study. **B.** Self-sampling kit consisting of swabs, transport tubes with solutions, instructions and biosecurity bag. **C.** Self-sampling instructions, which were provided together with a detailed briefing by a researcher and/or personnel from HR department of the organizations involved in the study.

A.

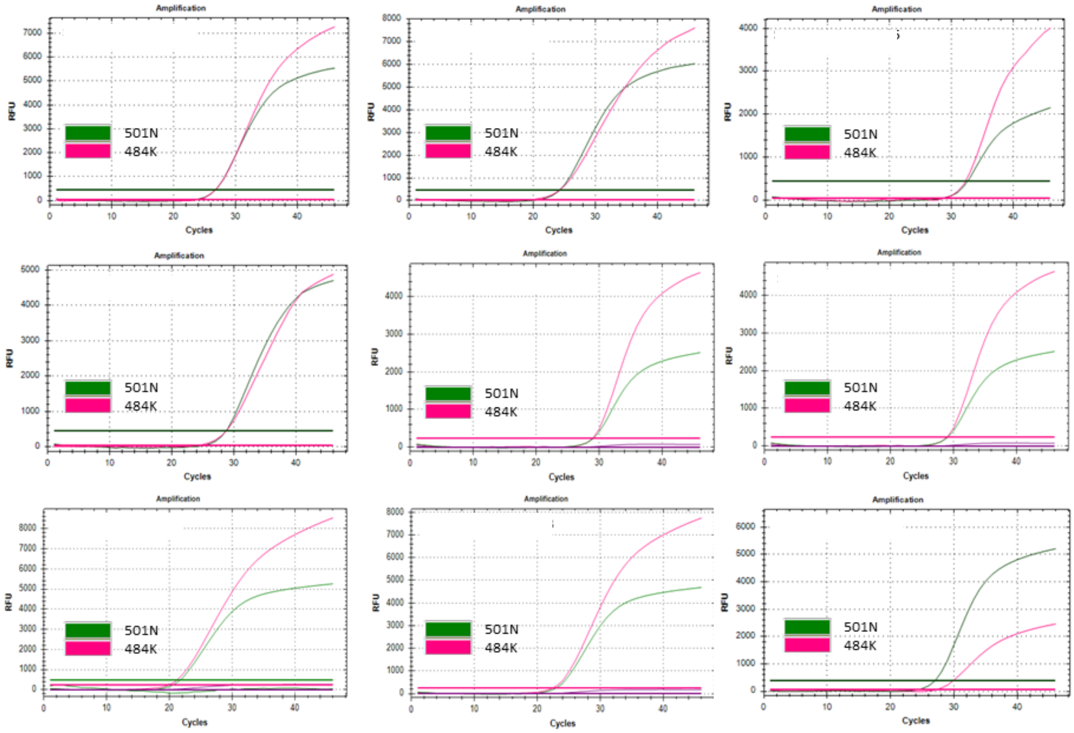


B.

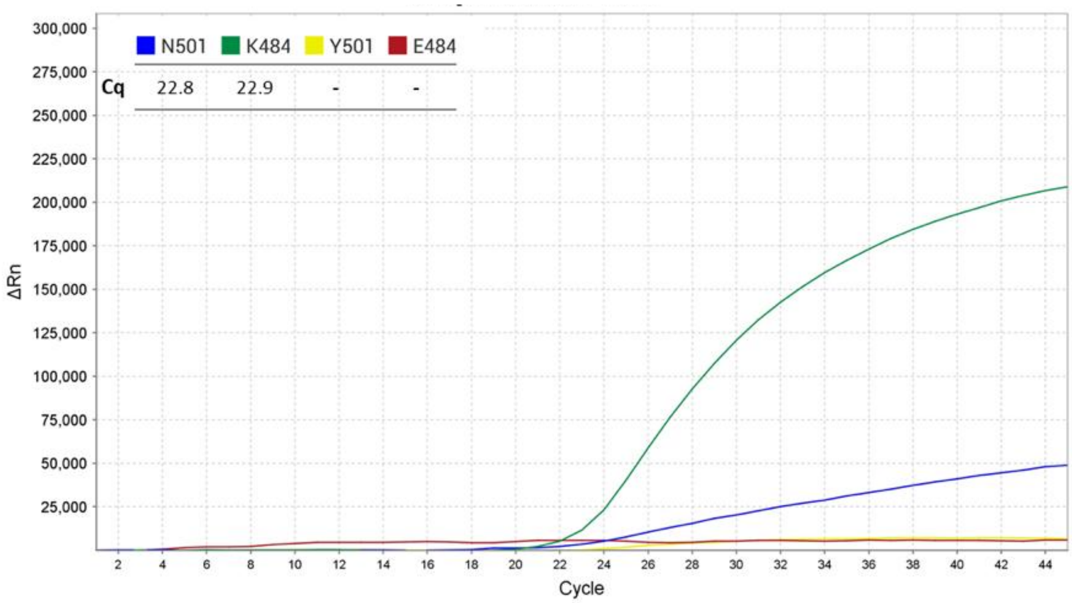


**Figure S2. RT-qPCR screening of E484K mutation in Jalisco (A) and San Luis Potosi (B).** The RT-qPCR curves corresponding to mutation E484K (red) and wild-type N501 sequence (green) are shown. All samples tested were previously diagnosed as positive for SARS-CoV-2. The detection assay is described in the Methods section. ID of each sample were removed to protect patients privacy.

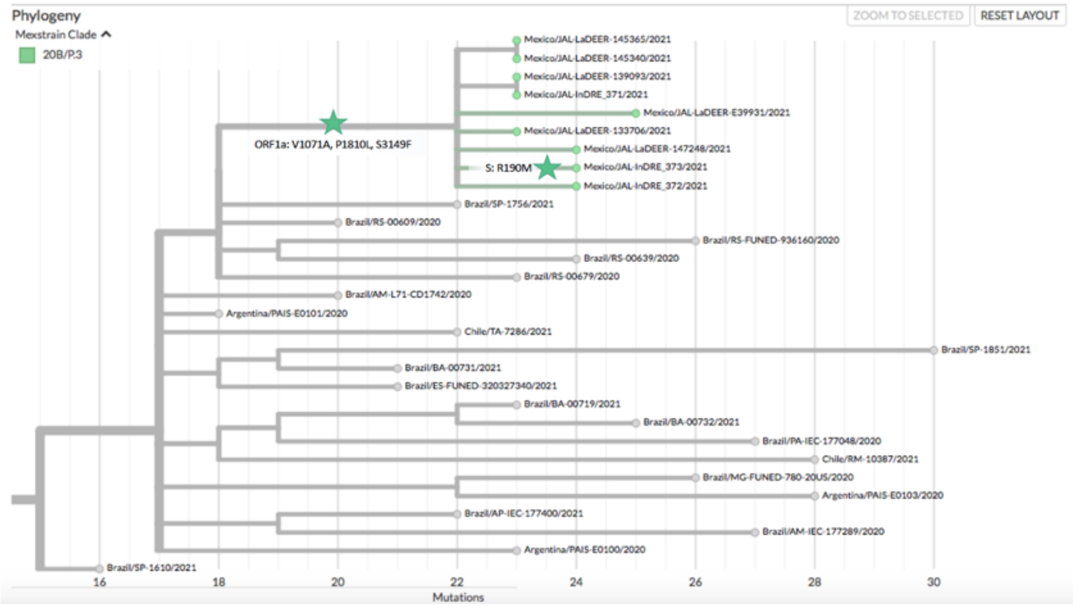


**Figure S3. P.3 distinctive sub-clade.** This VOI is supported by the mutations ORF1a: V1071A, P1810L, S3149F and S: R190M, shown here in more detail (see **Figure 1C**).

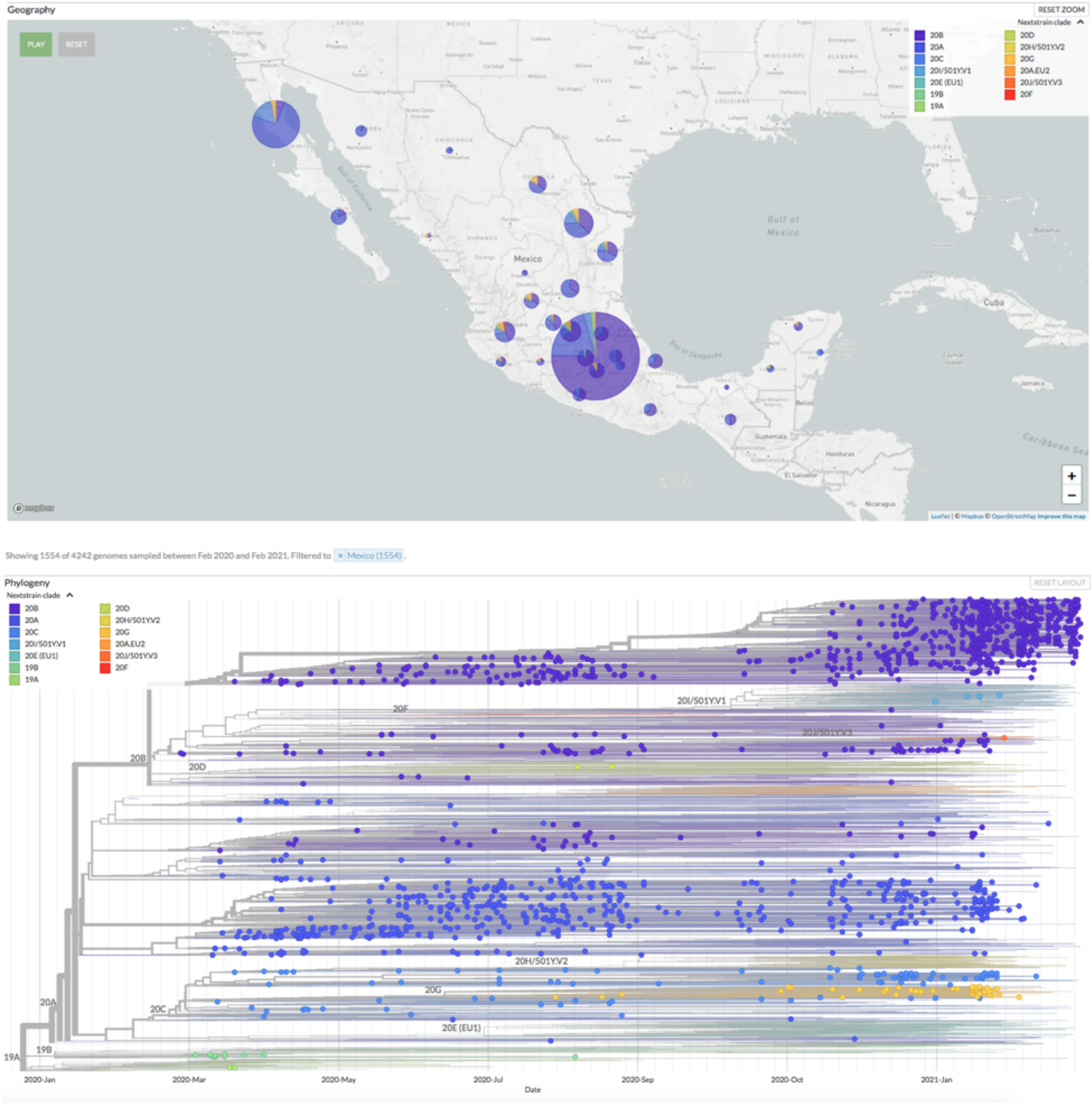


**Figure S4-a.** Nextstrain clades and geographic distribution in Mexico between March 2020 to February 2021.

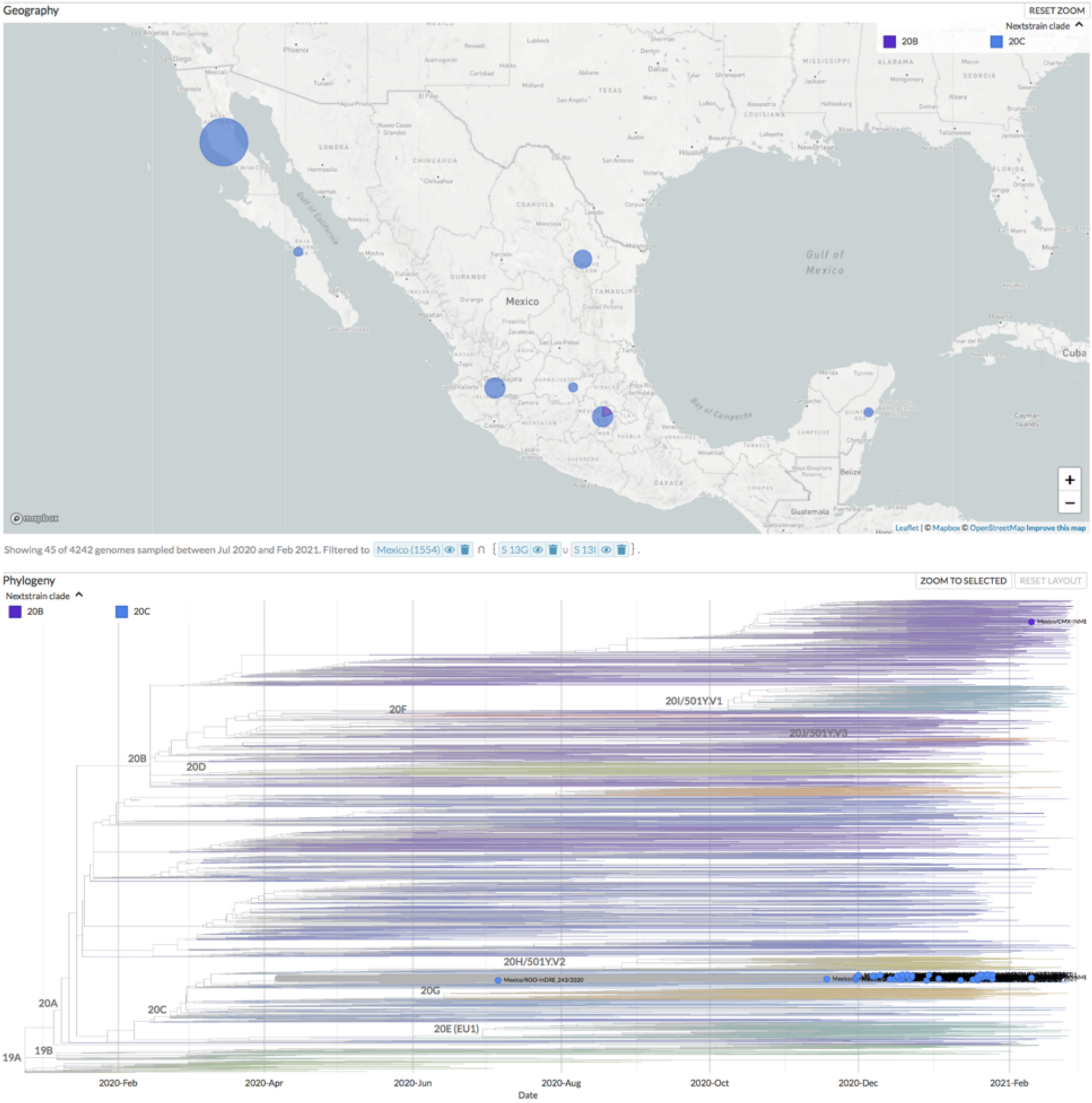


**Figure S4-b**. Nextstrain clades and geographic distribution in Mexico between February 2020 to February 2021 showing the **mutation Spike: 13G/I**.

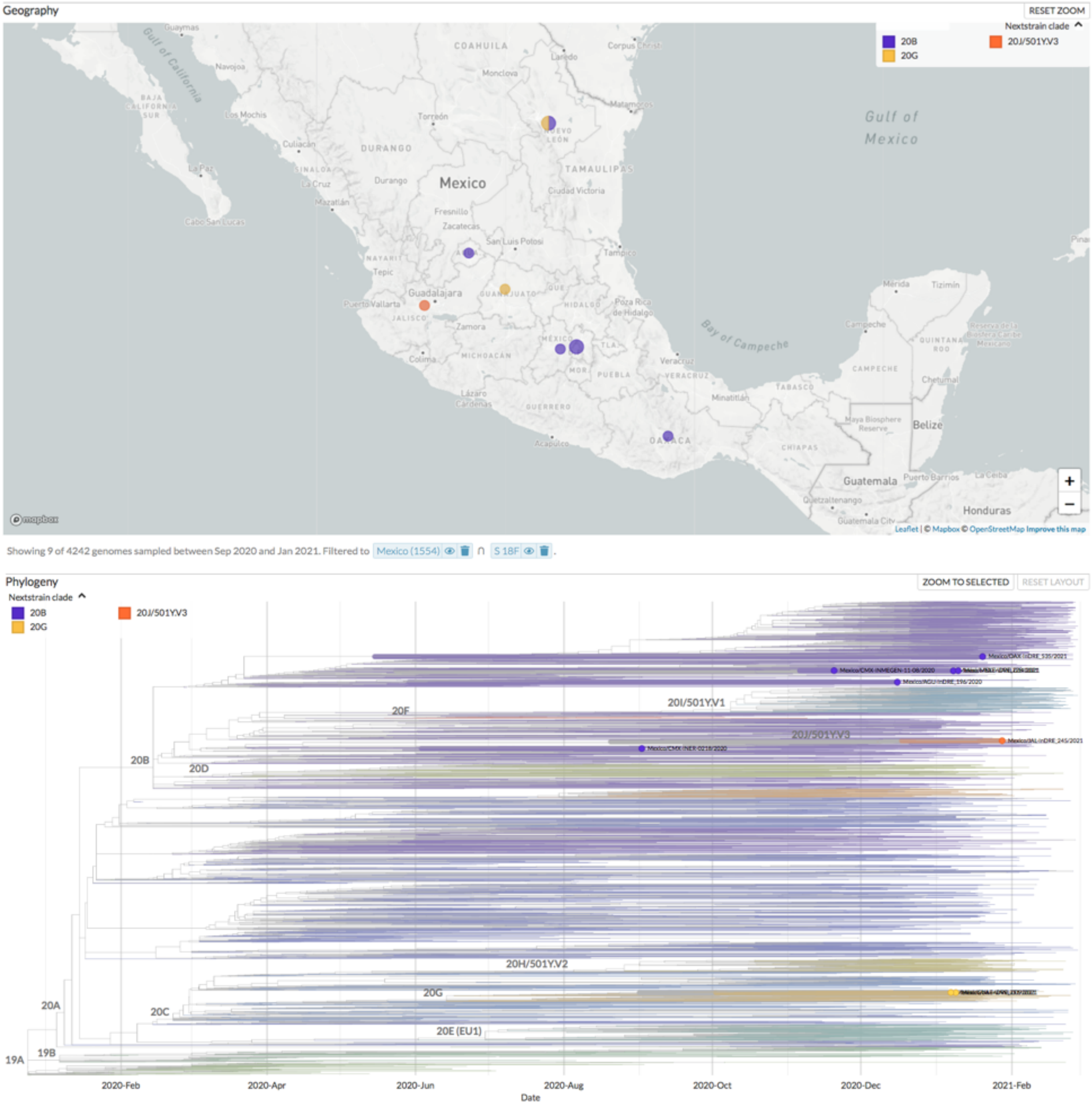


**Figure S4-c.** Nextstrain clades and geographic distribution in Mexico between February 2020 to February 2021 showing the mutation **Spike: 18F**.

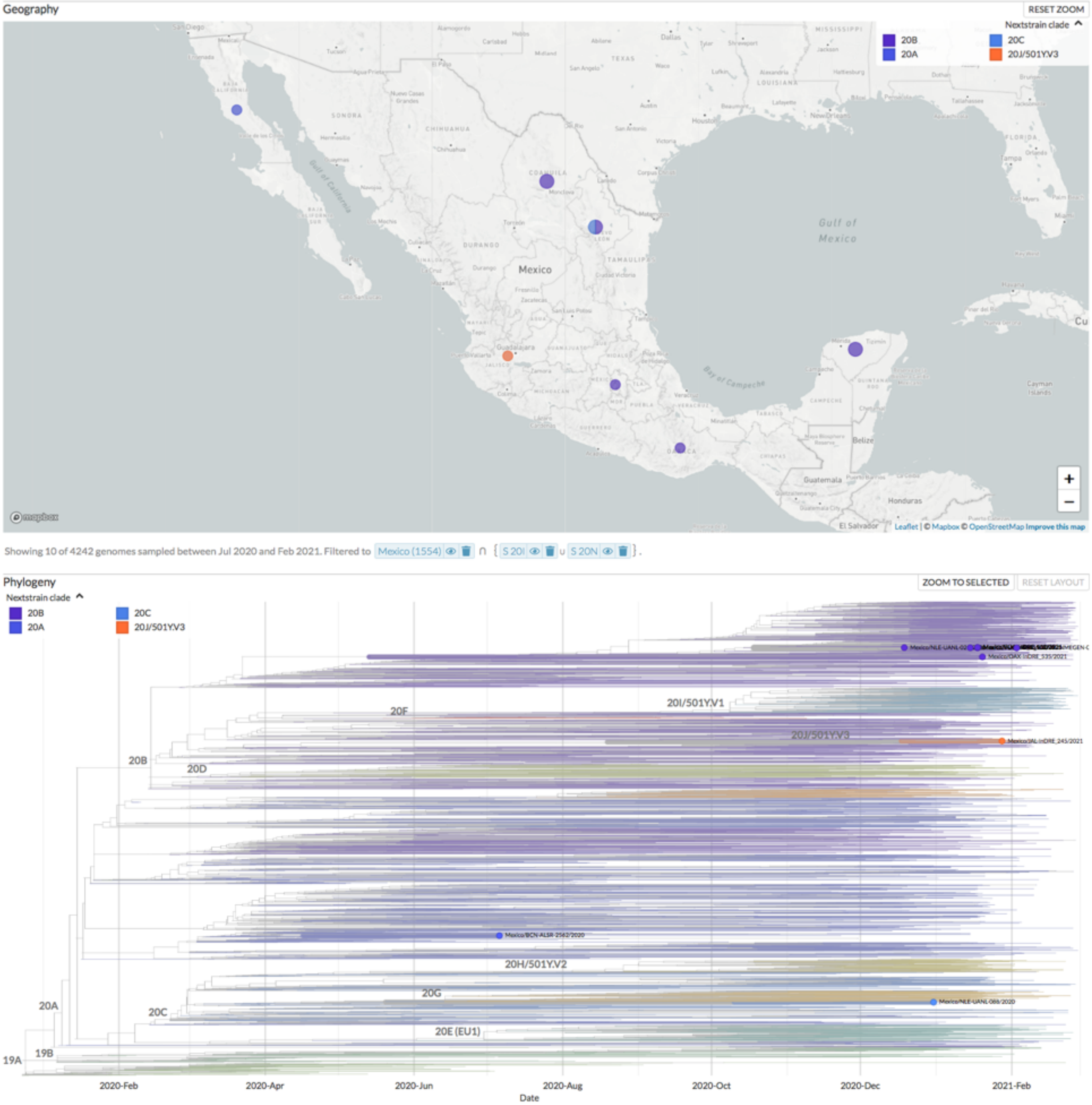


**Figure S4-d.** Nextstrain clades and geographic distribution in Mexico between February 2020 to February 2021 showing the mutation **Spike: 20N/I**.

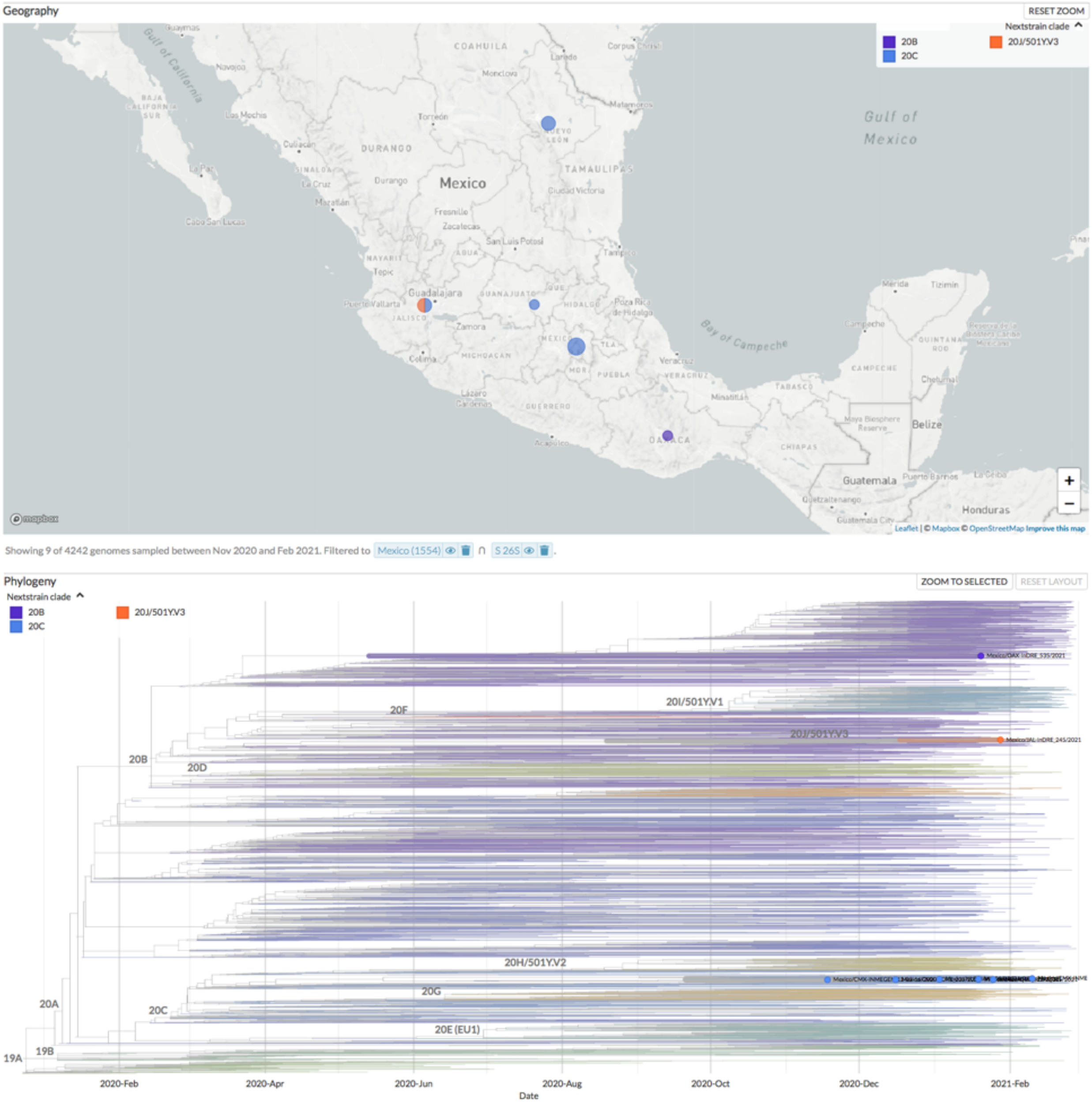


**Figure S4-e.** Nextstrain clades and geographic distribution in Mexico between February 2020 to February 2021 showing the mutation **Spike: 26S**.

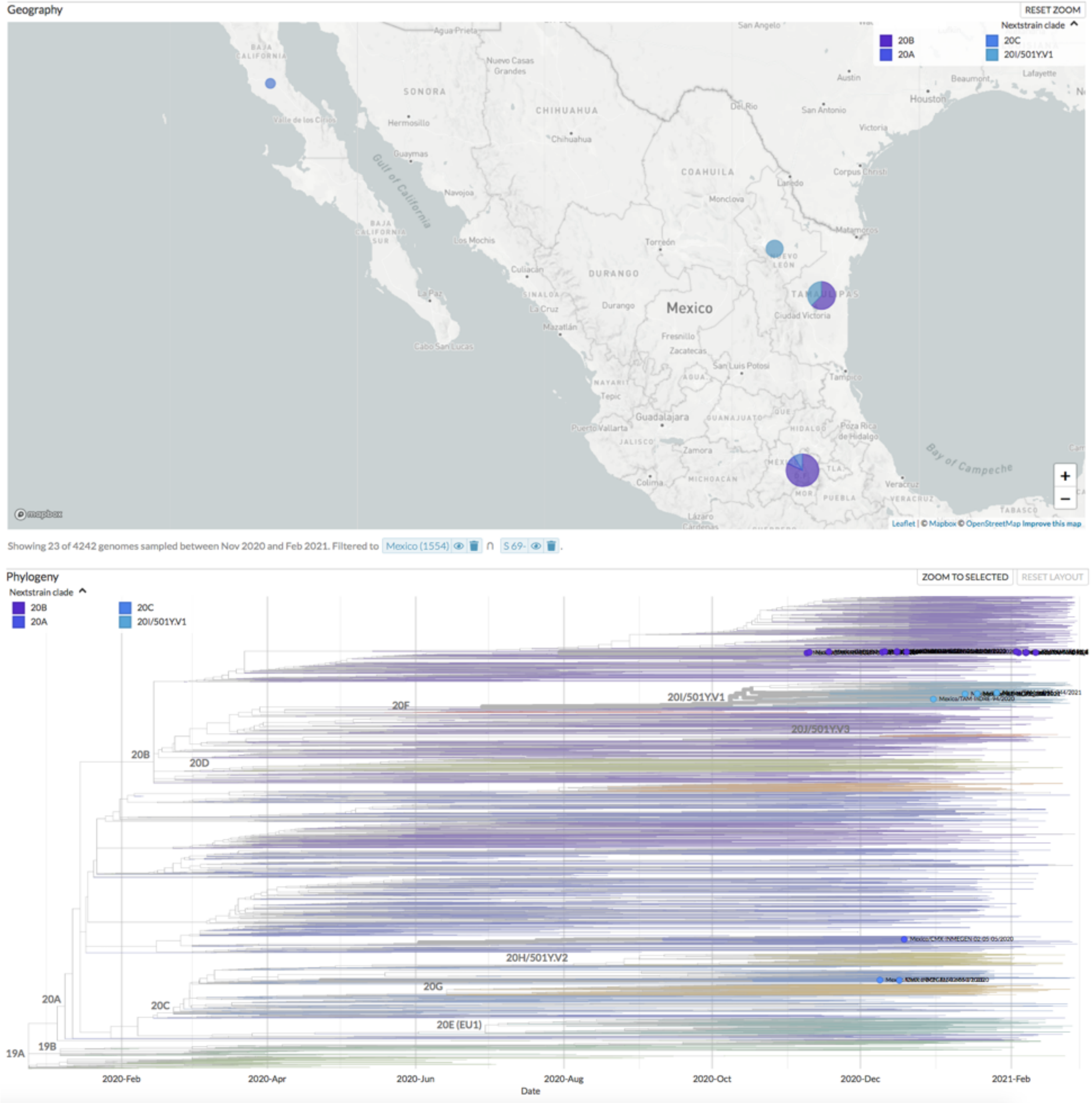


**Figure S4-f.** Nextstrain clades and geographic distribution in Mexico between February 2020 to February 2021 showing the mutation **Spike: 69X-70X (deletion)**.

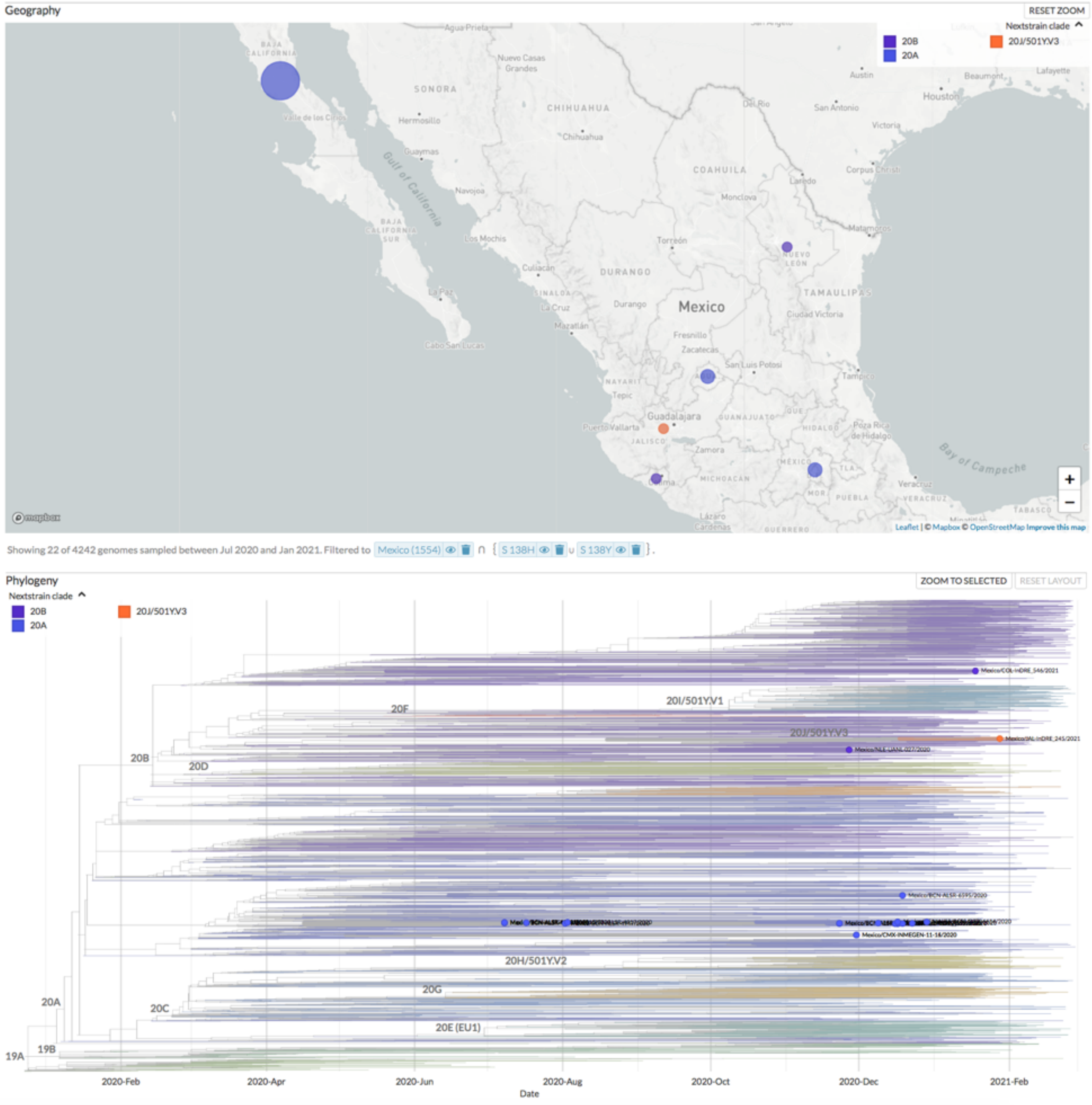


**Figure S4-g.** Nextstrain clades and geographic distribution in Mexico between February 2020 to February 2021 showing the mutation **Spike: 138Y/H**.

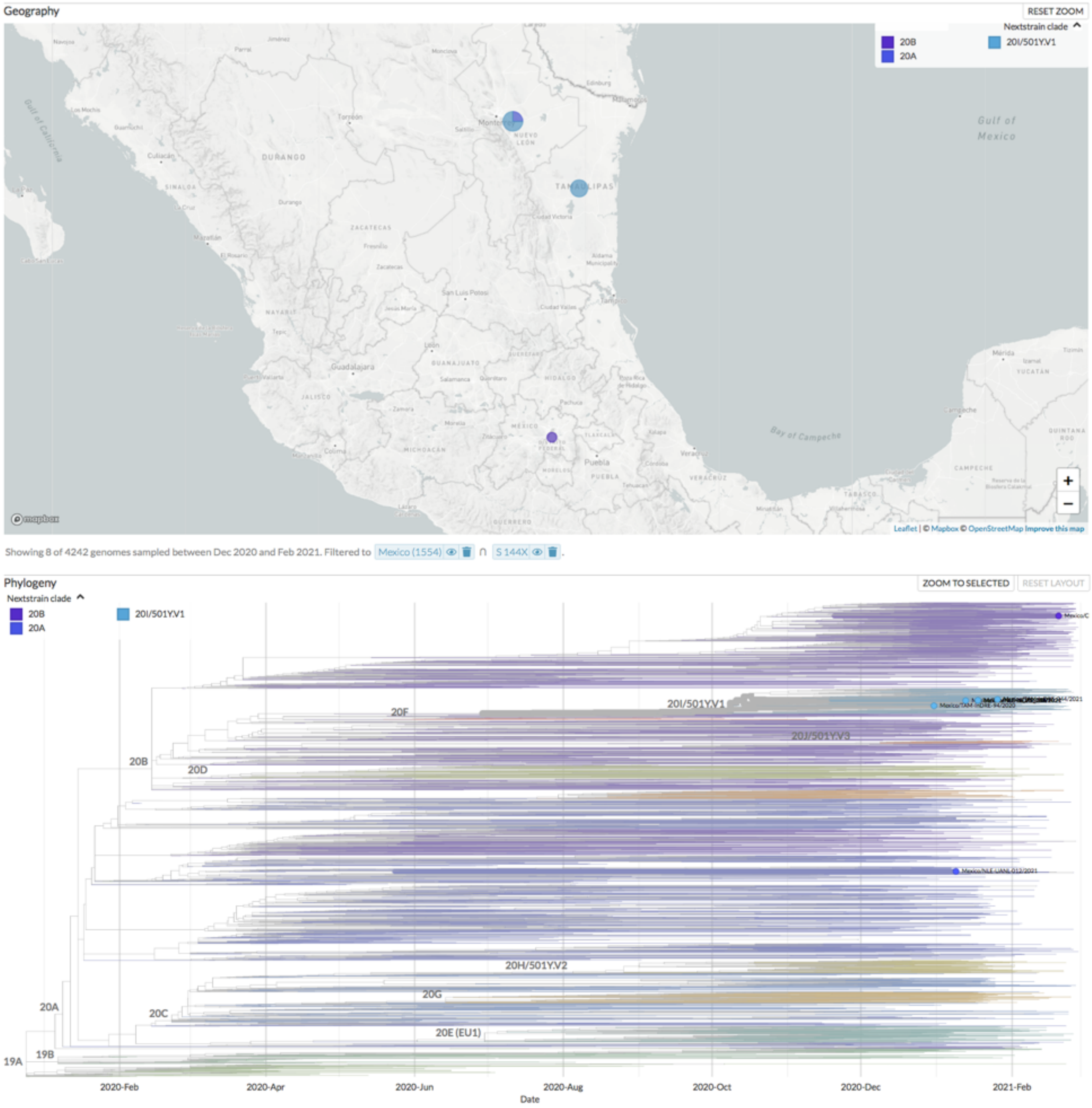


**Figure S4-h.** Nextstrain clades and geographic distribution in Mexico between February 2020 to February 2021 showing the mutation **Spike: 144X (deletion)**.

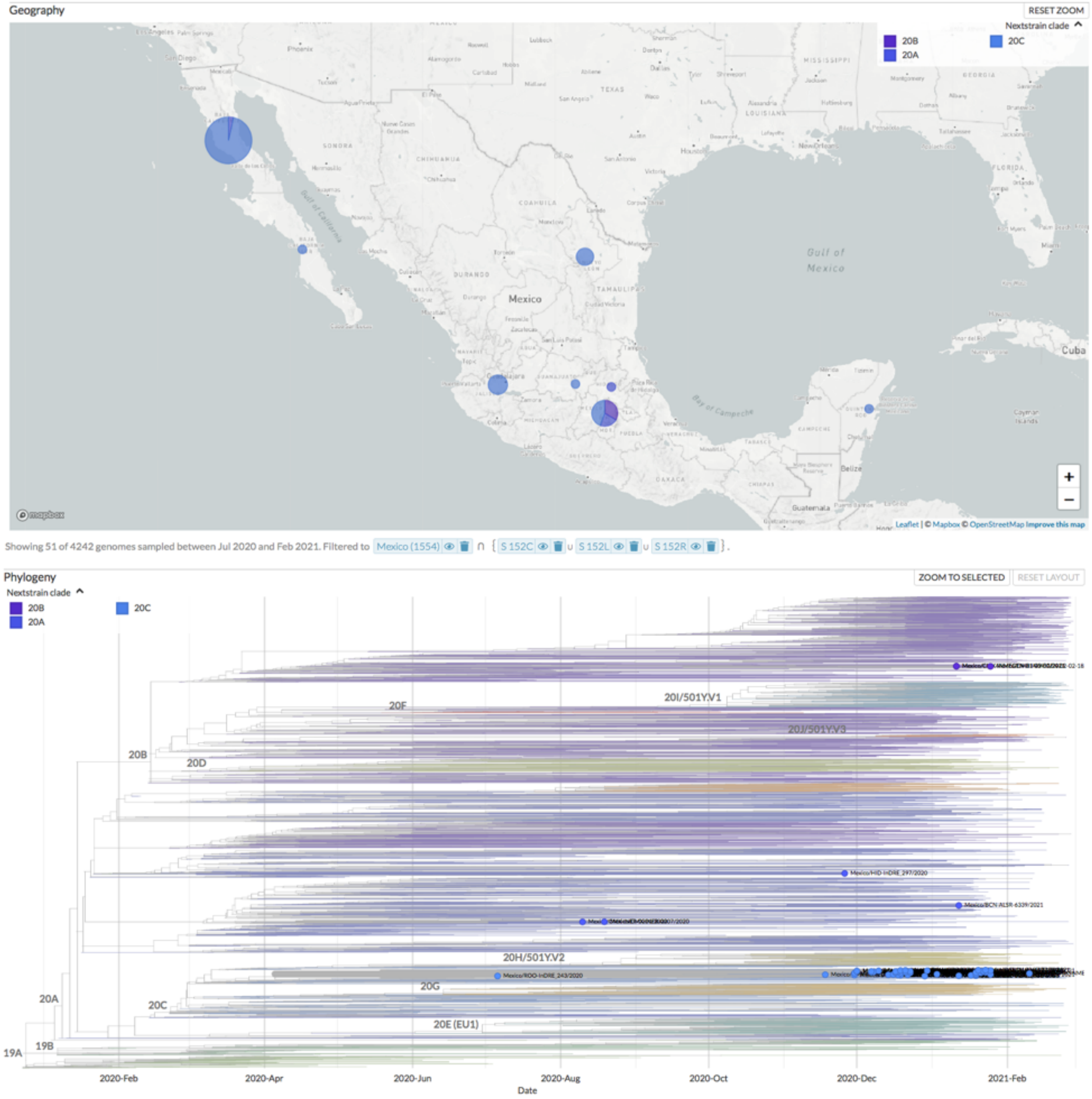


**Figure S4-i.** Nextstrain clades and geographic distribution in Mexico between February 2020 to February 2021 showing the mutation **Spike: 152C/L/R.**

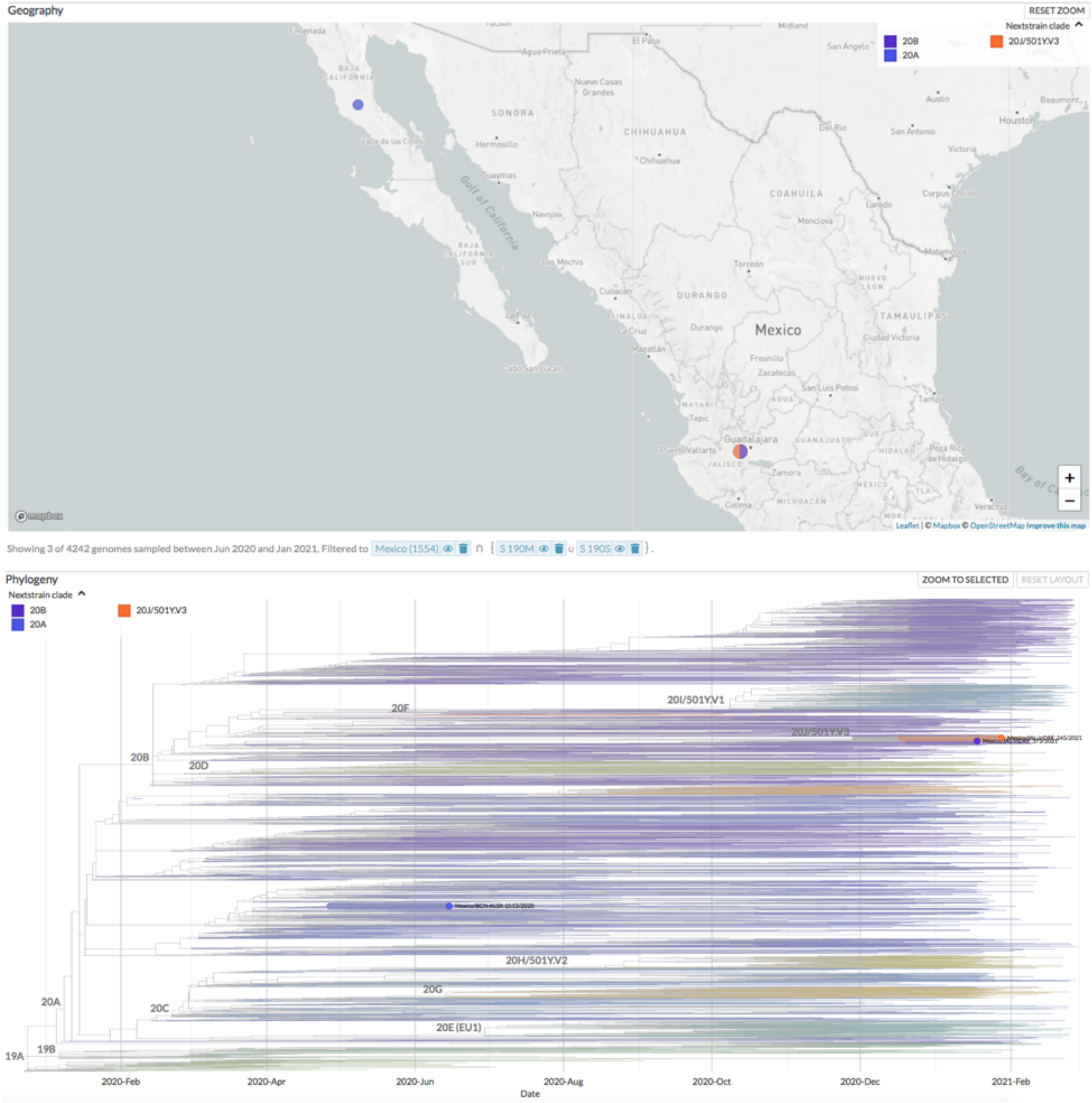


**Figure S4-j.** Nextstrain clades and geographic distribution in Mexico between February 2020 to February 2021 showing the mutation **Spike: 190M/S**.

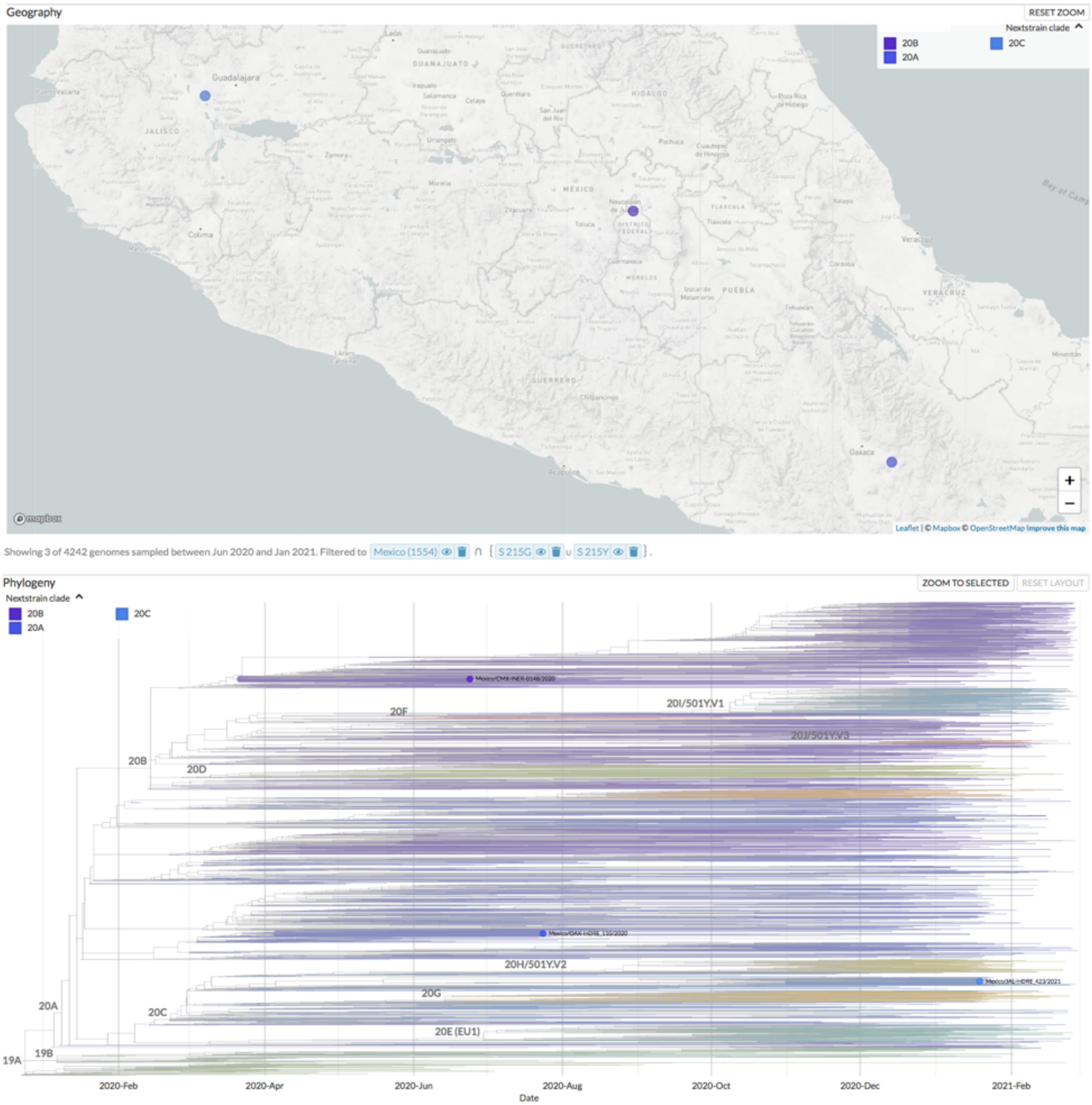


**Figure S4-k.** Nextstrain clades and geographic distribution in Mexico between February 2020 to February 2021 showing the mutation **Spike: 215G/Y**.

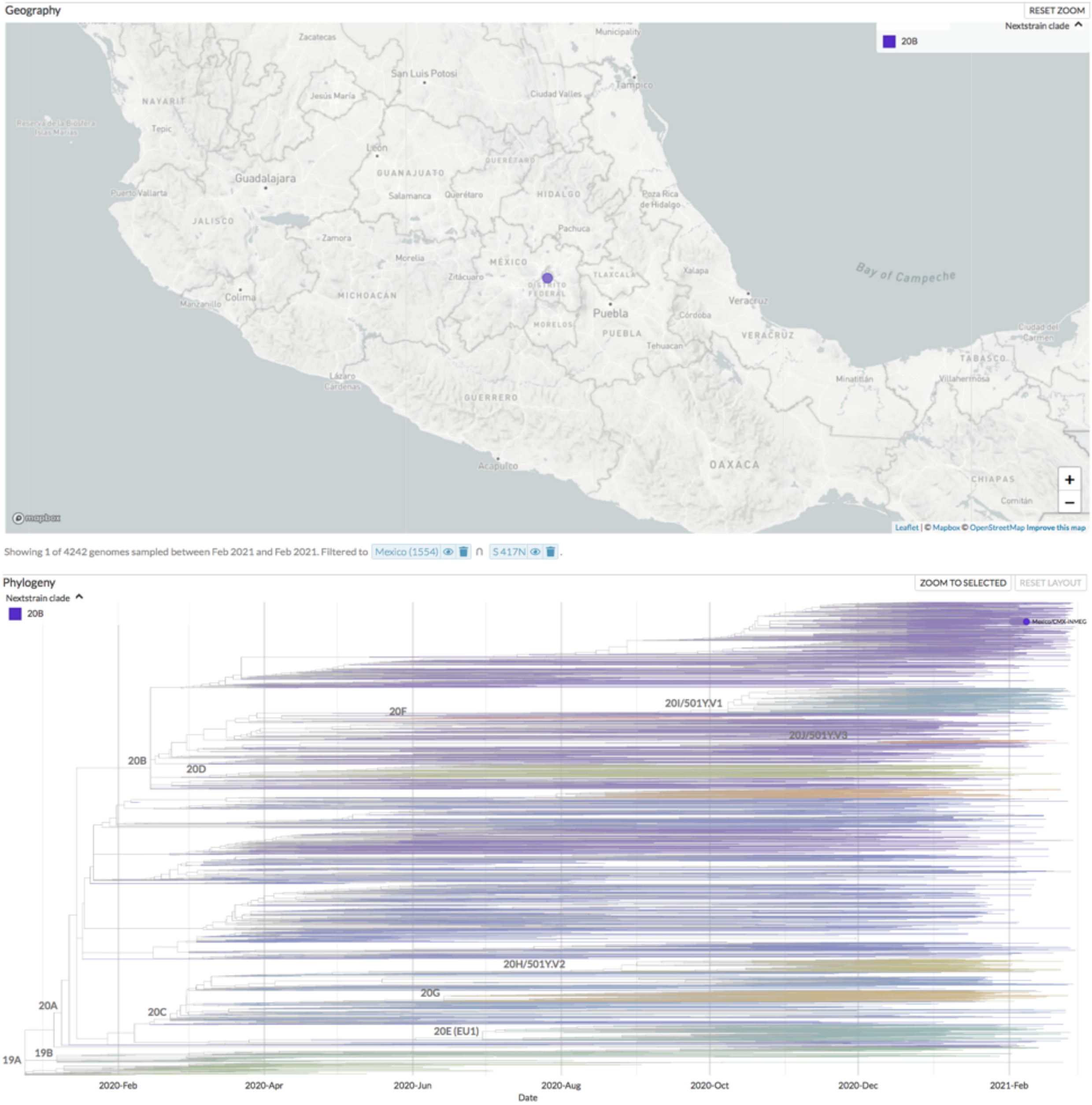


**Figure S4-l.** Nextstrain clades and geographic distribution in Mexico between February 2020 to February 2021 showing the mutation **Spike: 417N**.

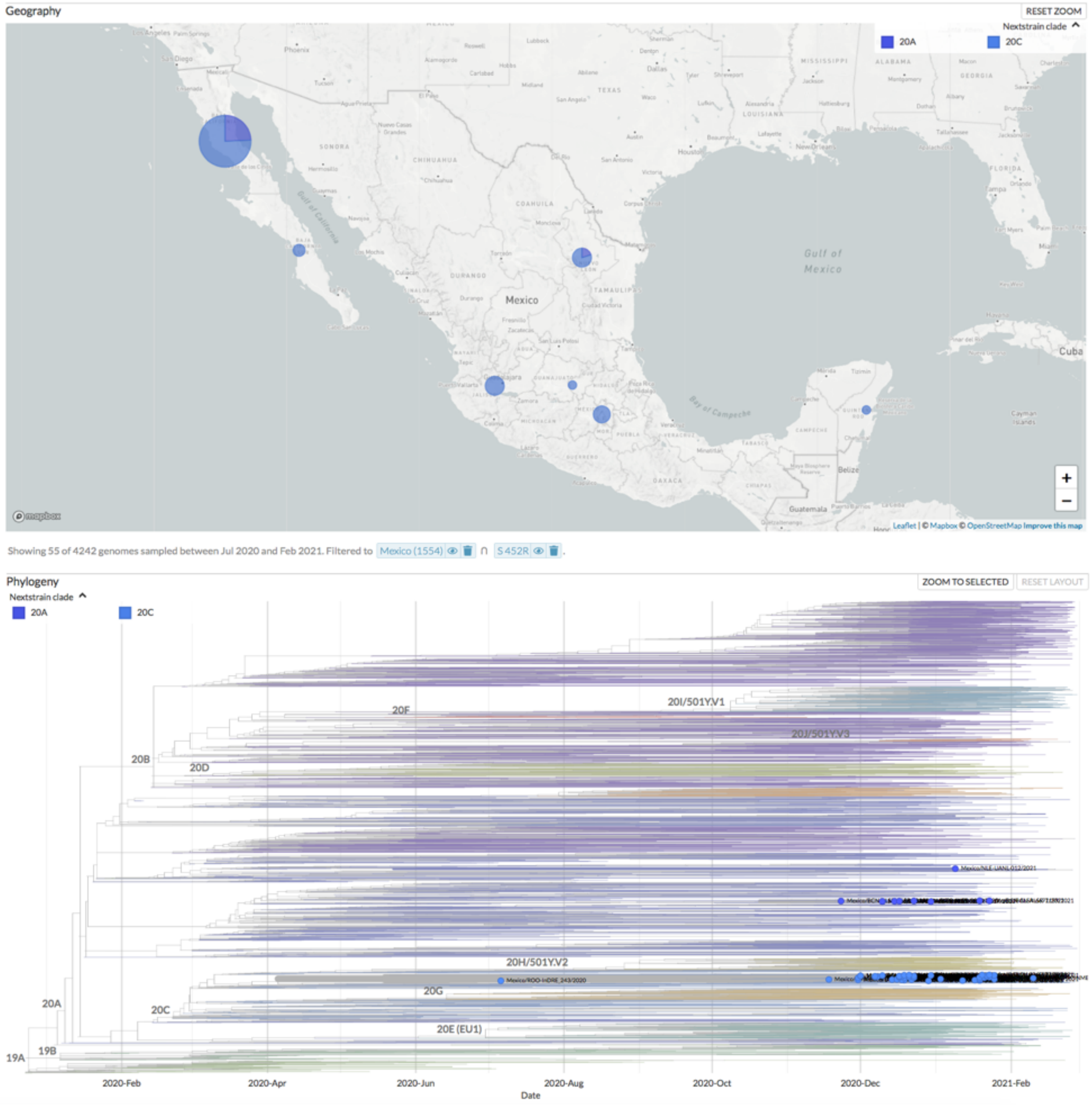


**Figure S4-m**. Nextstrain clades and geographic distribution in Mexico between February 2020 to February 2021 showing the mutation **Spike: 452R**.

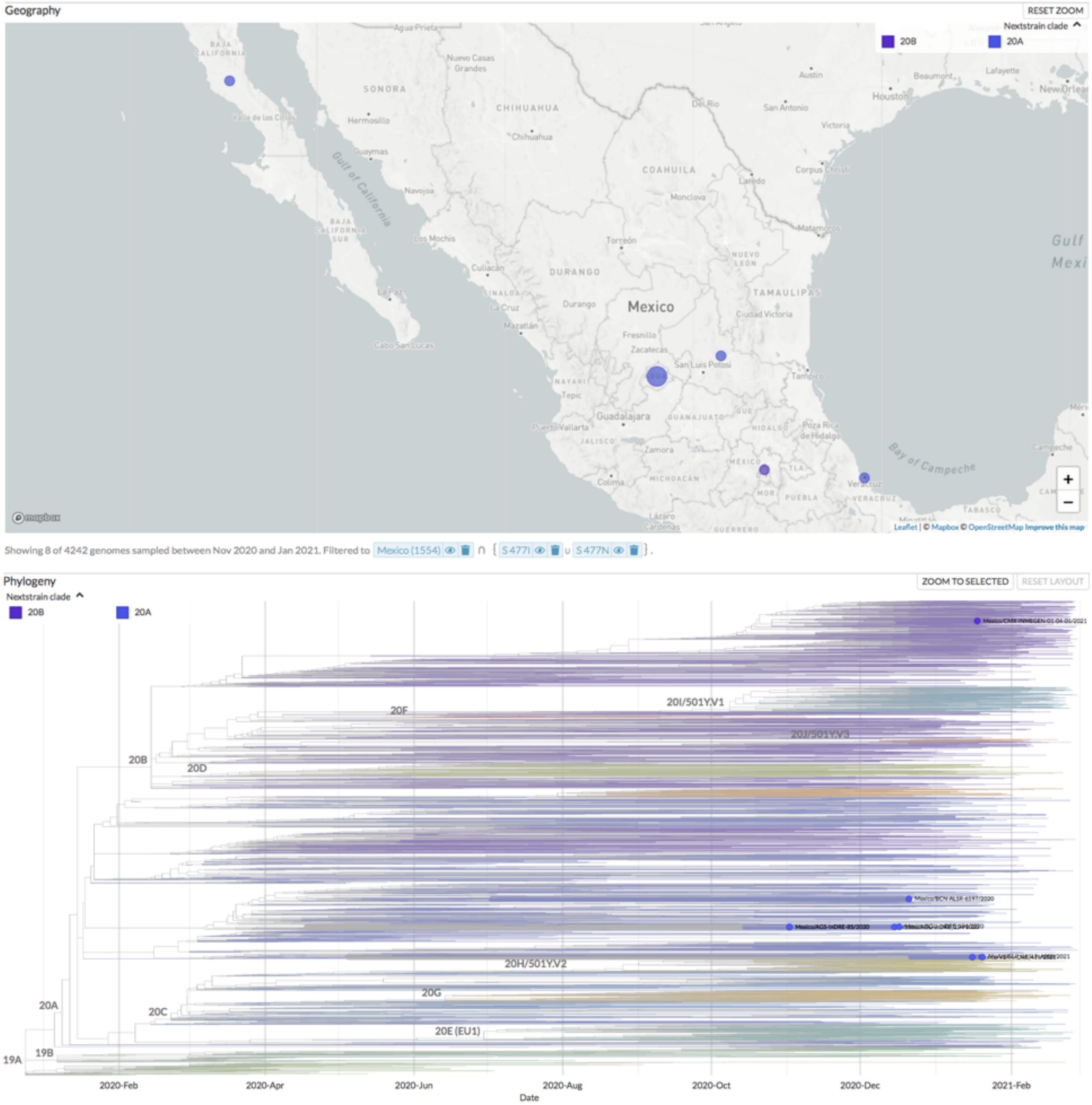


**Figure S4-n.** Nextstrain clades and geographic distribution in Mexico between February 2020 to February 2021 showing the mutation Spike: **477N/I.**

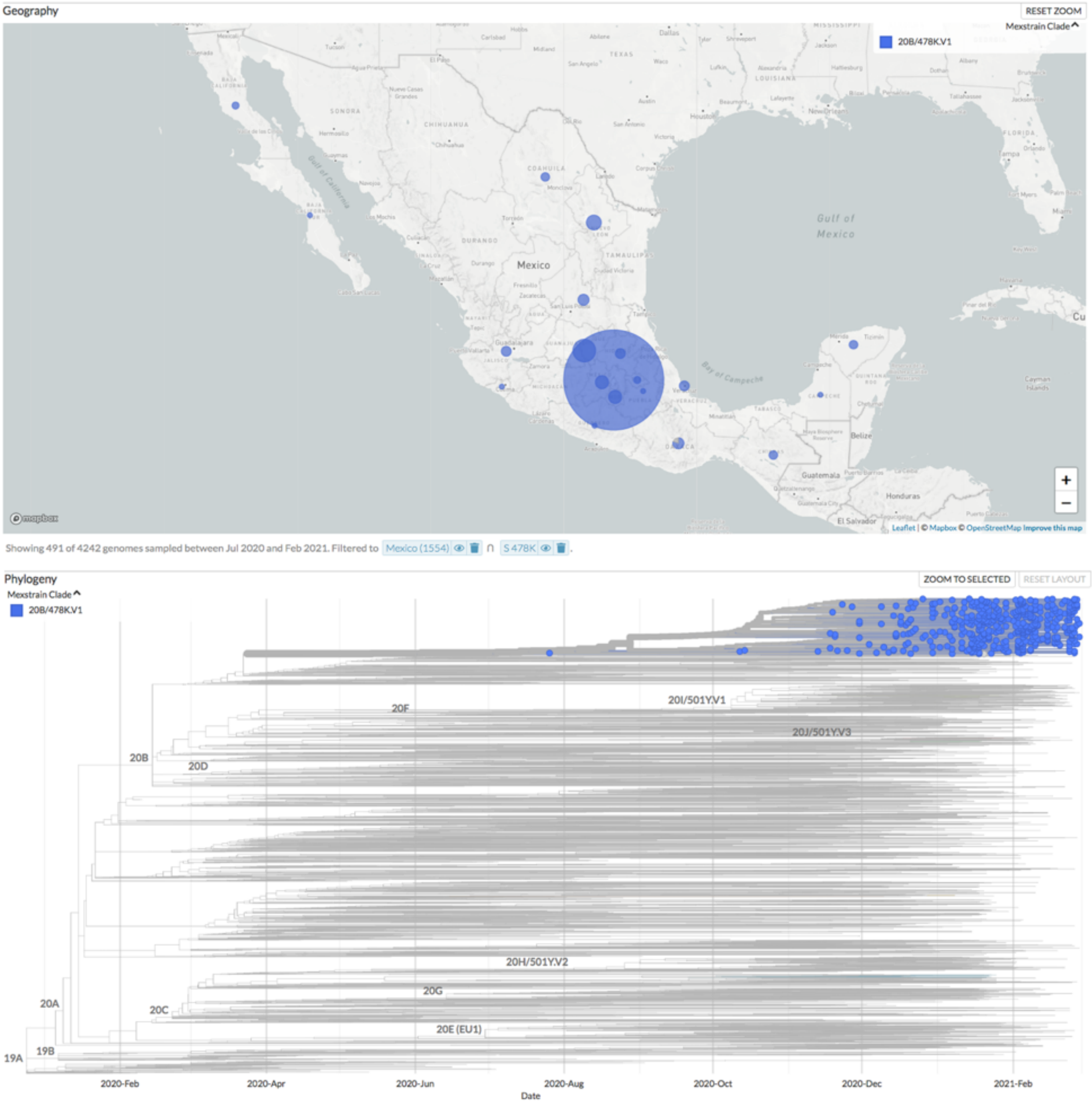


**Figure S4-o.** Nextstrain clades and geographic distribution in Mexico between February 2020 to February 2021 showing the mutation **Spike: 478K**.

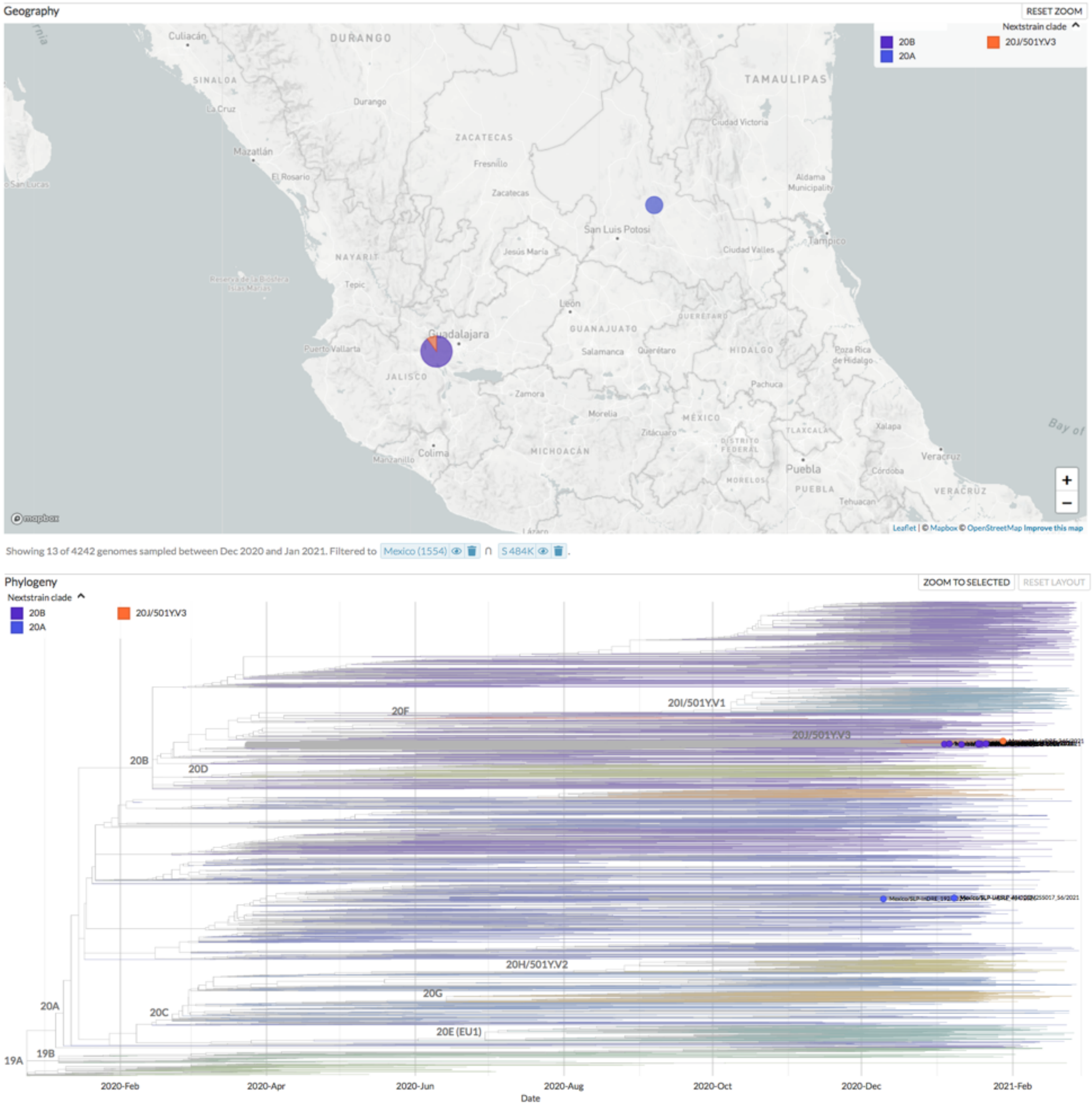


**Figure S4-p.** Nextstrain clades and geographic distribution in Mexico between February 2020 to February 2021 showing the mutation **Spike: 484K**.

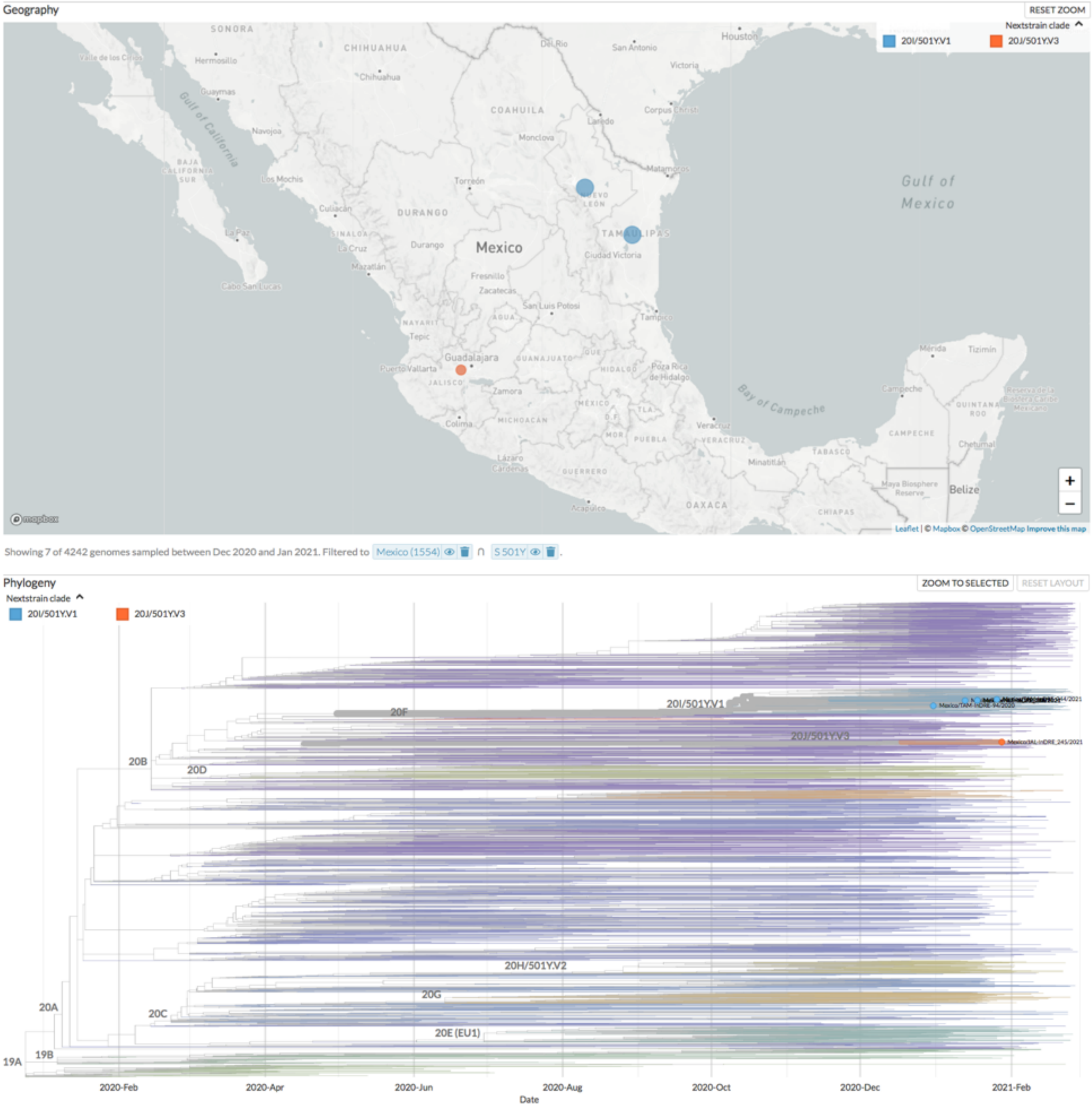


**Figure S4-q.** Nextstrain clades and geographic distribution in Mexico between February 2020 to February 2021 showing the mutation **Spike: 501Y**.

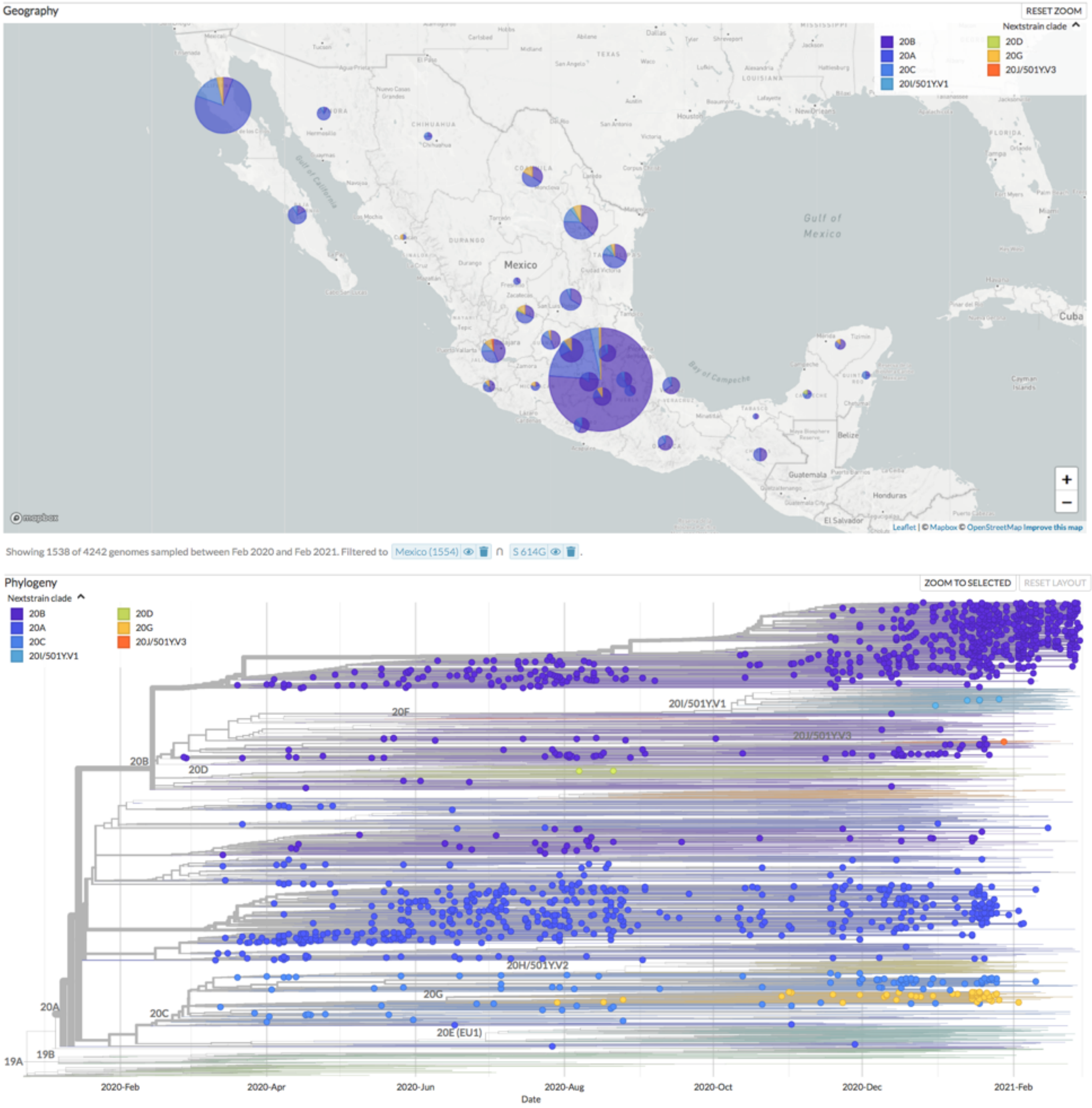


**Figure S4-r.** Nextstrain clades and geographic distribution in Mexico between February 2020 to February 2021 showing the mutation **Spike: 614G**.

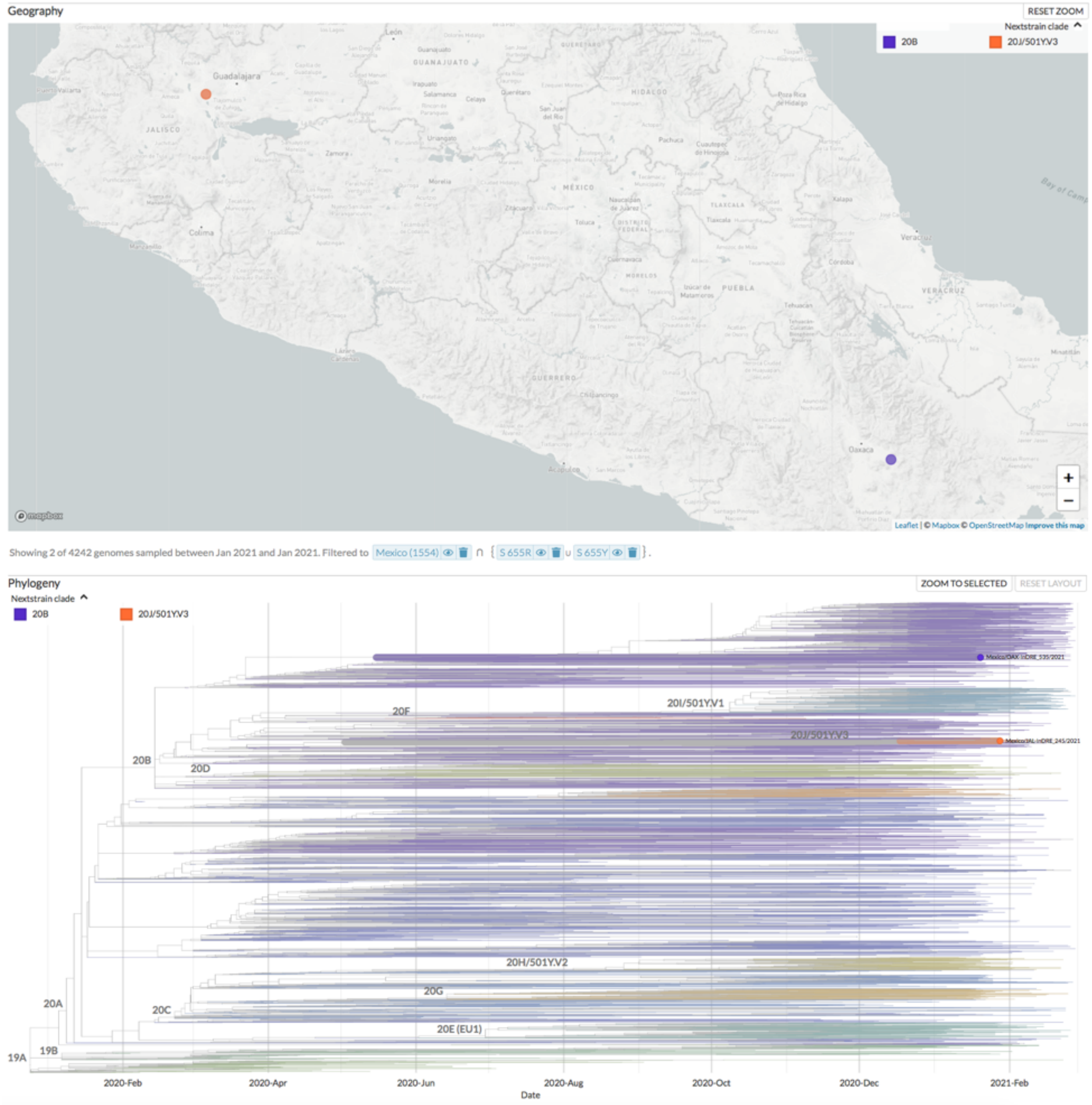


**Figure S4-s**. Nextstrain clades and geographic distribution in Mexico between February 2020 to February 2021 showing the mutation **Spike: 655Y/R**.

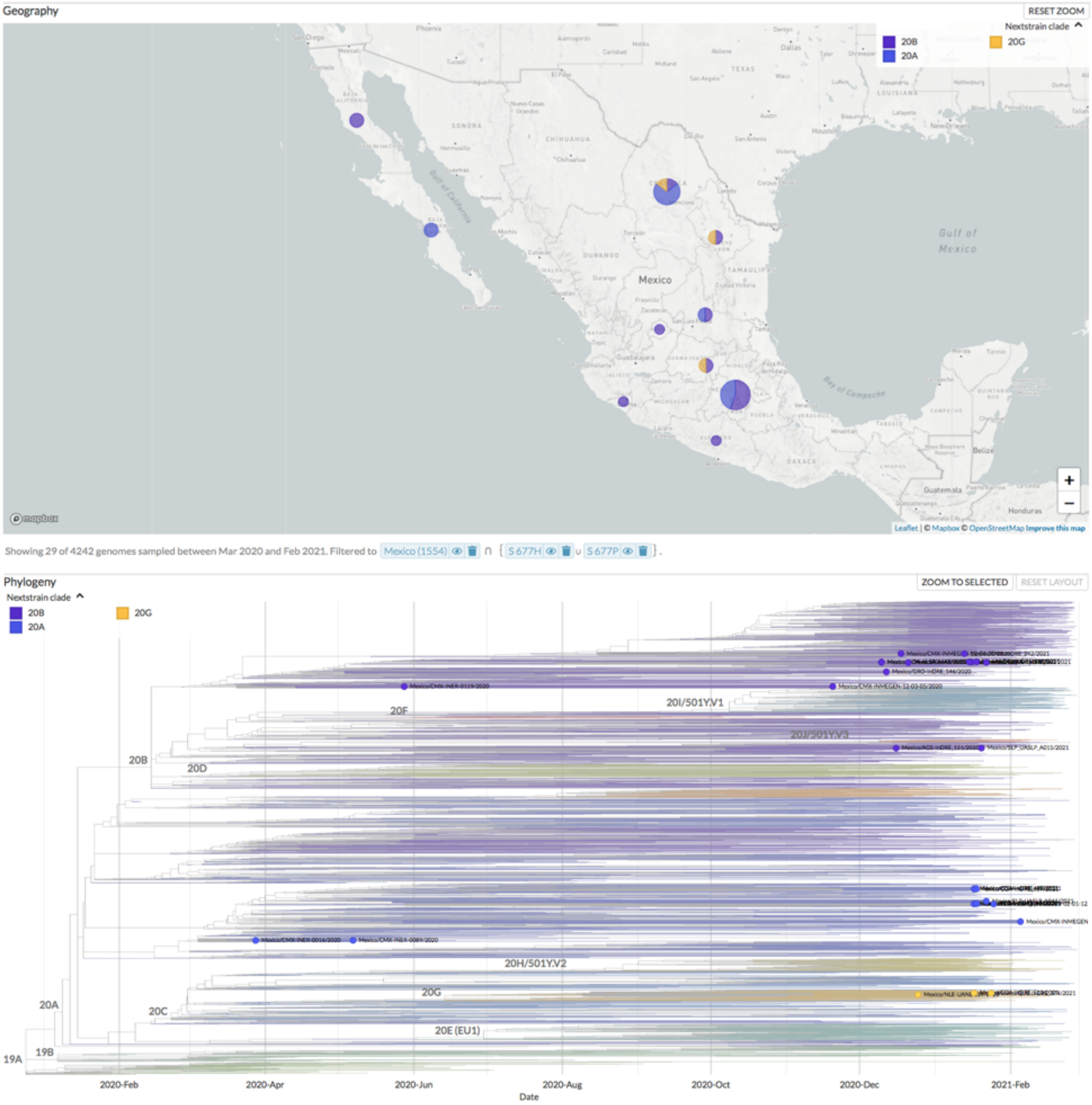


**Figure S4-t.** Nextstrain clades and geographic distribution in Mexico between February 2020 to February 2021 showing the mutation **Spike: 677H/P**.

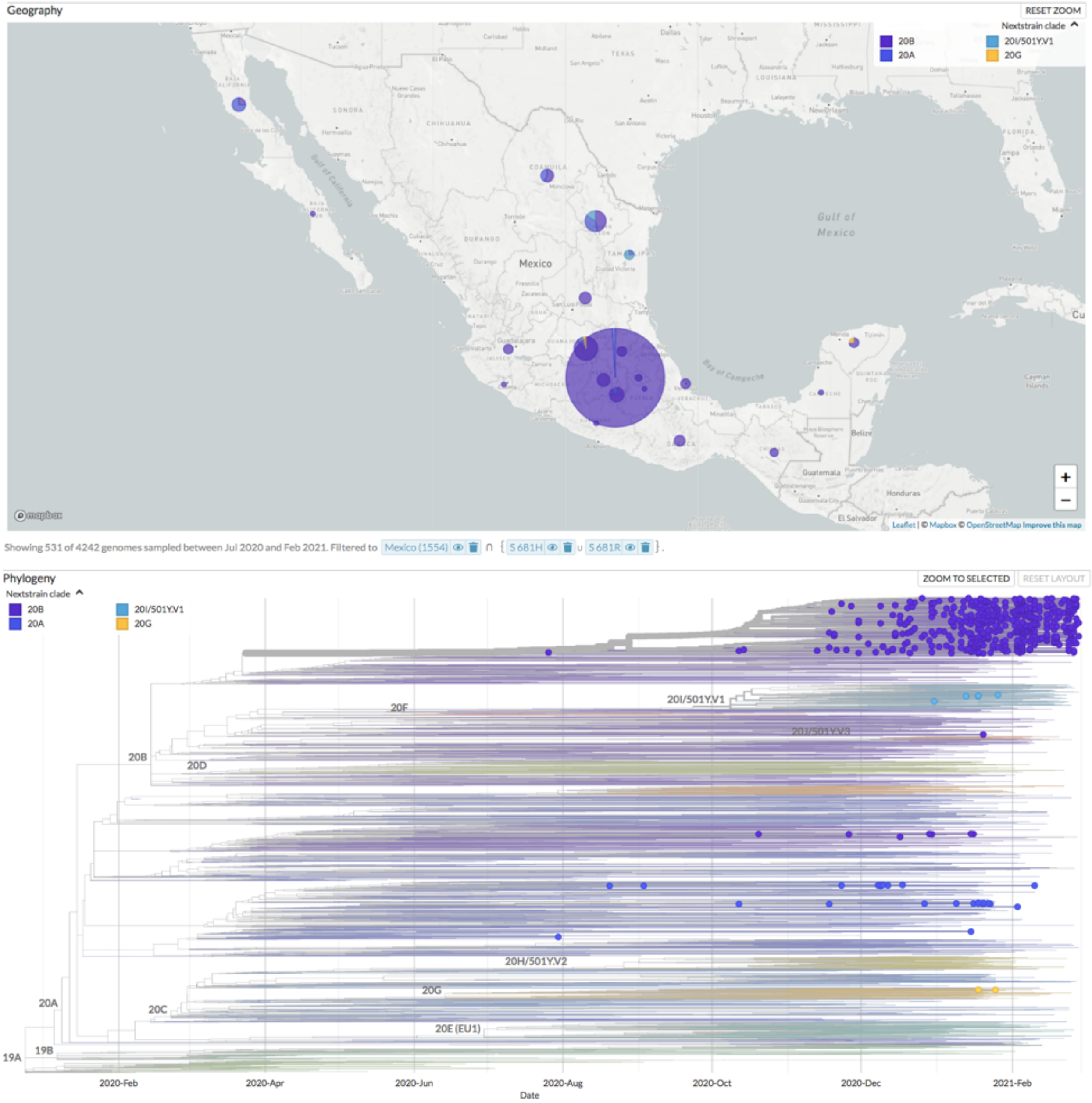


**Figure S4-u**. Nextstrain clades and geographic distribution in Mexico between February 2020 to February 2021 showing the mutation **Spike: 681H/R**.

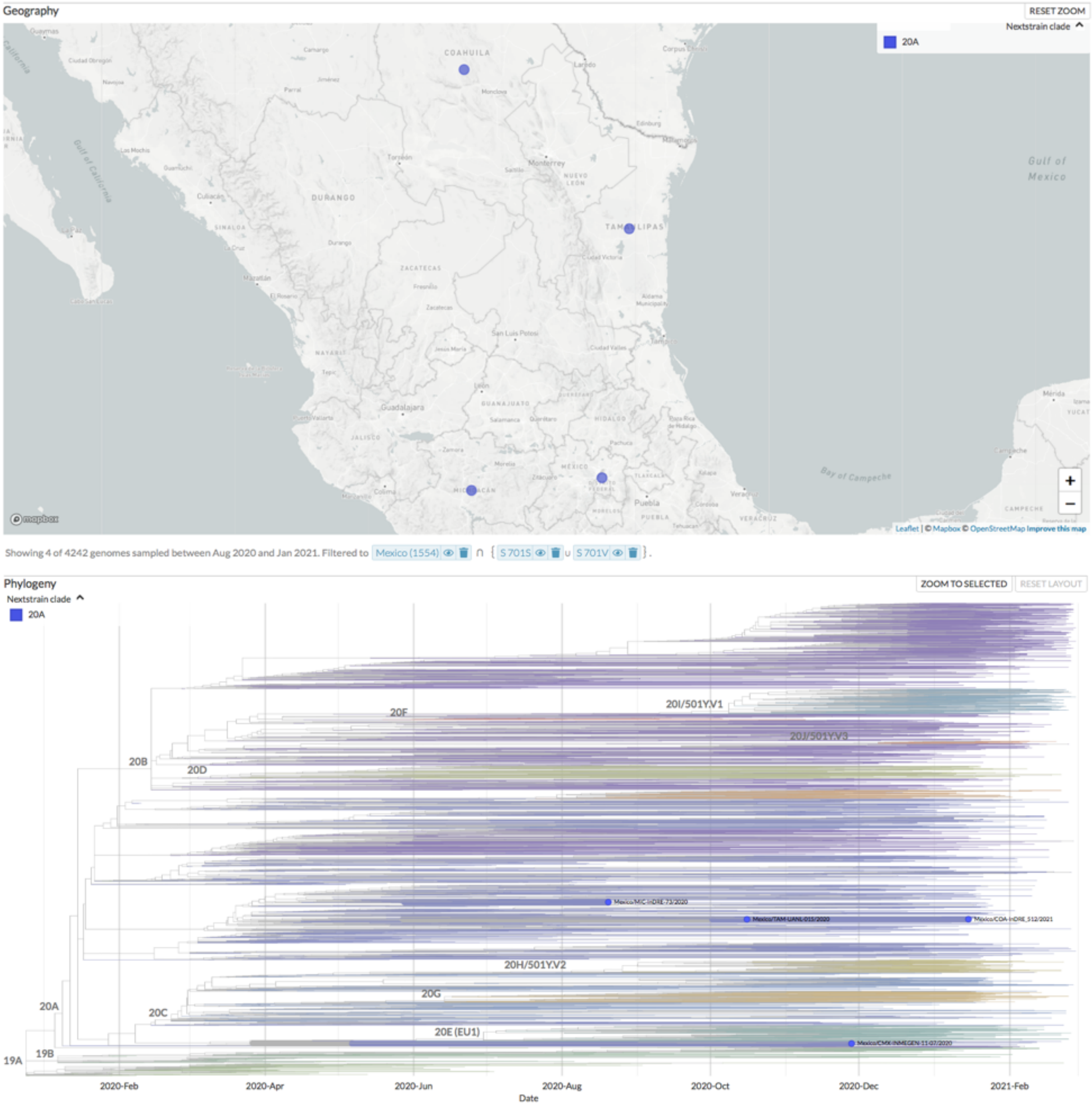


**Figure S4-v**. Nextstrain clades and geographic distribution in Mexico between February 2020 to February 2021 showing the mutation **Spike: 701S/V**.

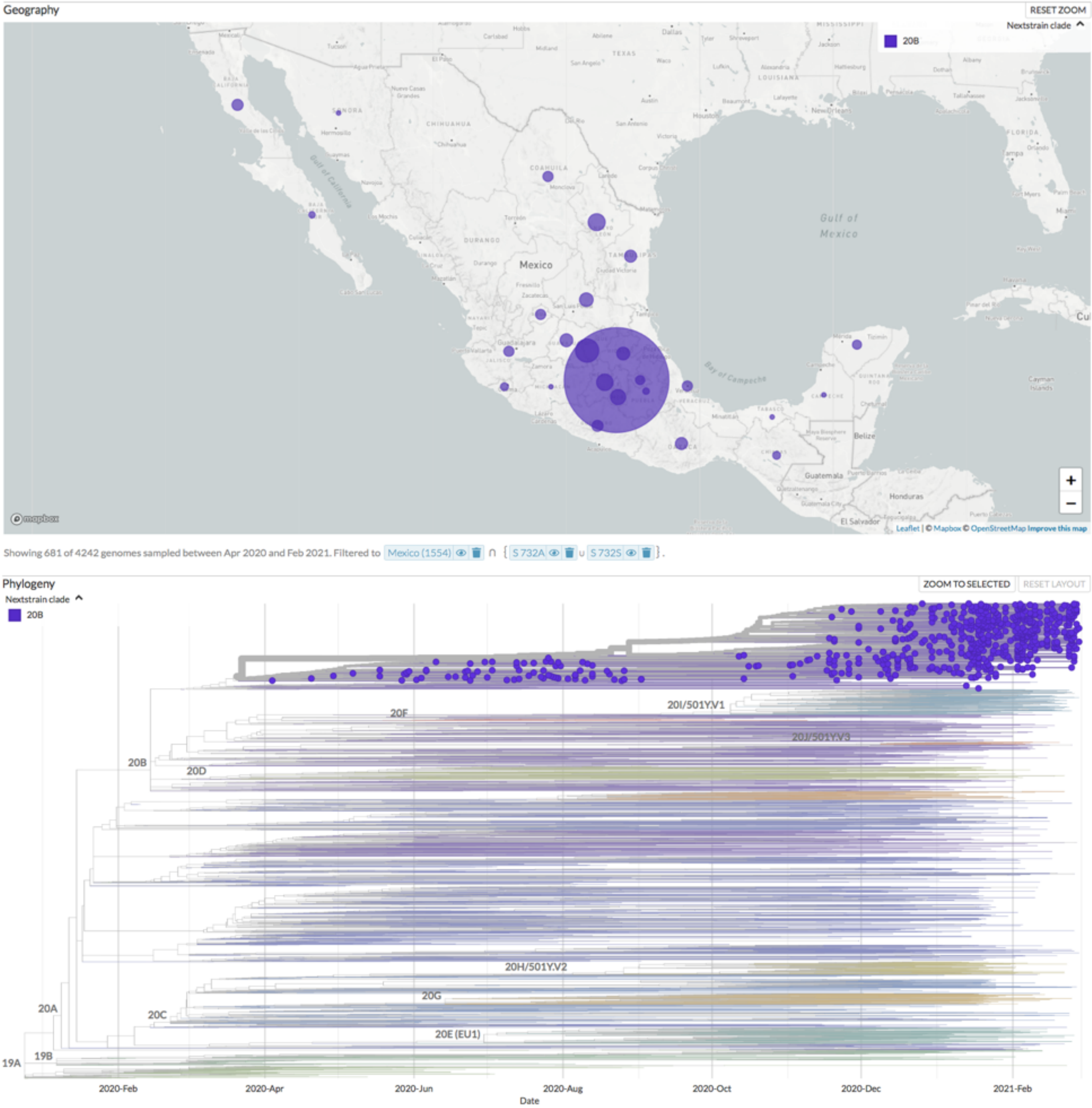


**Figure S4-w.** Nextstrain clades and geographic distribution in Mexico between February 2020 to February 2021 showing the mutation **Spike: 732A/S**.

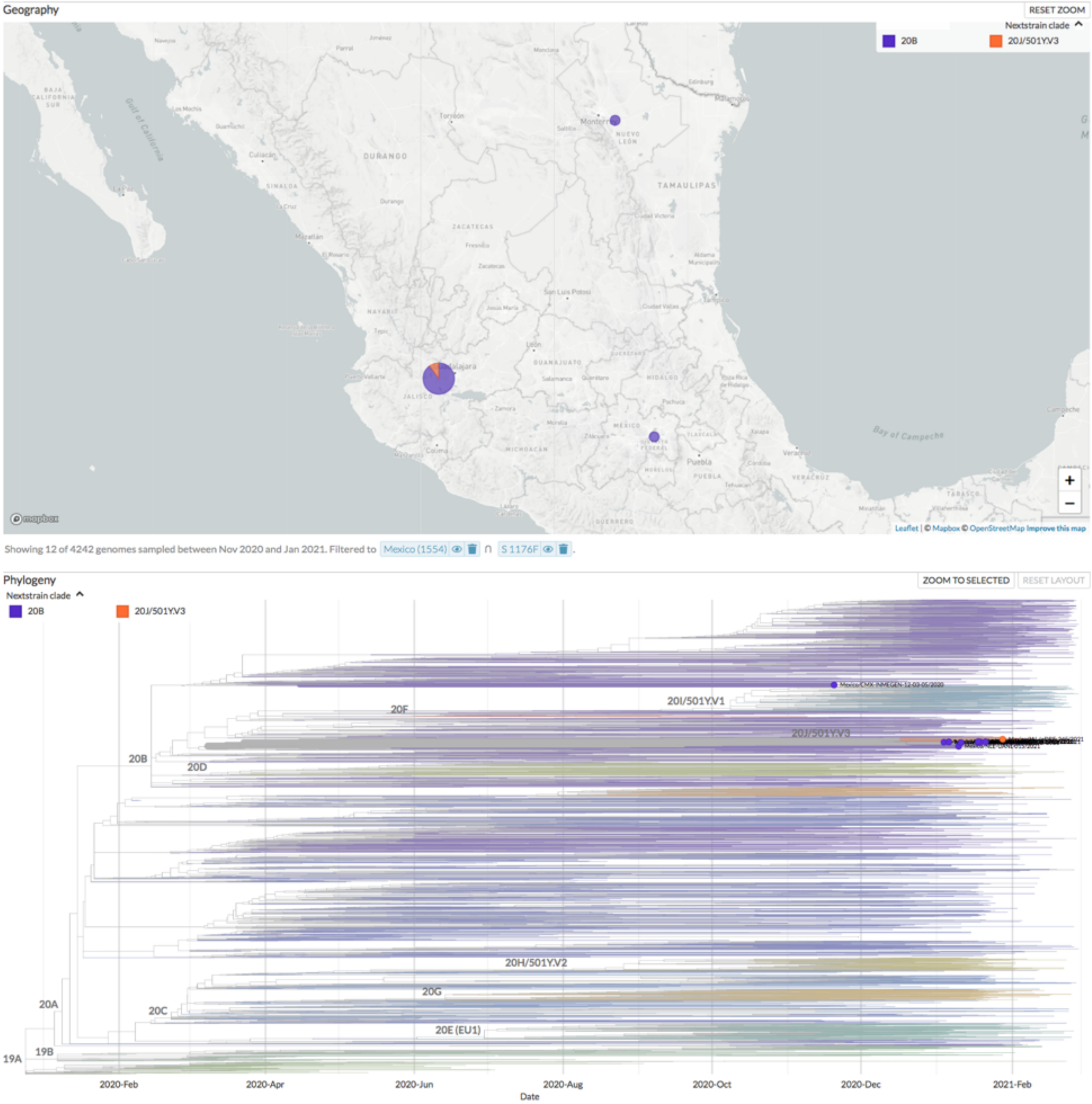


**Figure S4-x.** Nextstrain clades and geographic distribution in Mexico between February 2020 to February 2021 showing the mutation **Spike: 1176F.**

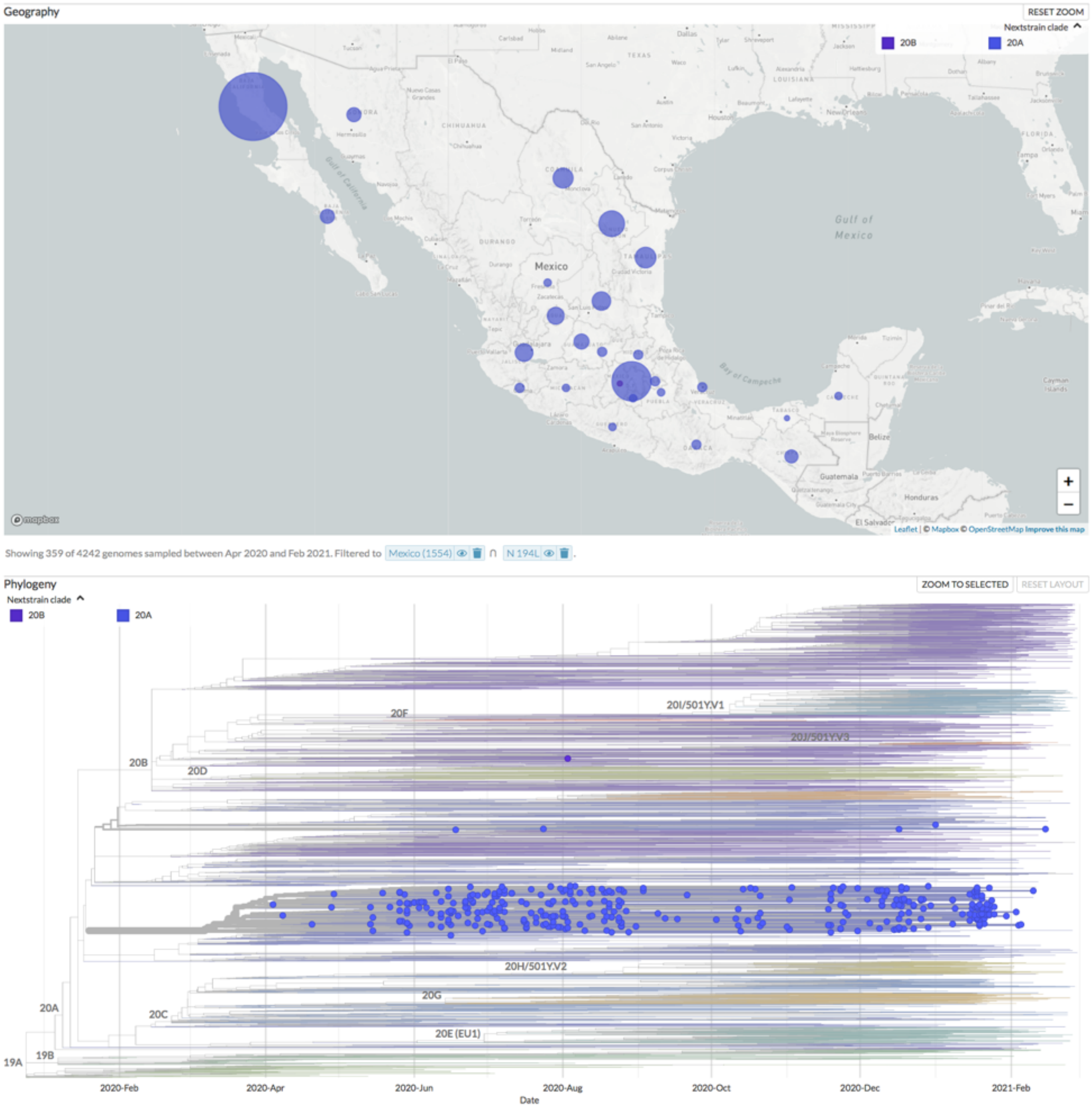


**Figure S4-y**. Nextstrain clades and geographic distribution in Mexico between February 2020 to February 2021 showing the mutation **Nucleocapsid: 194L.**

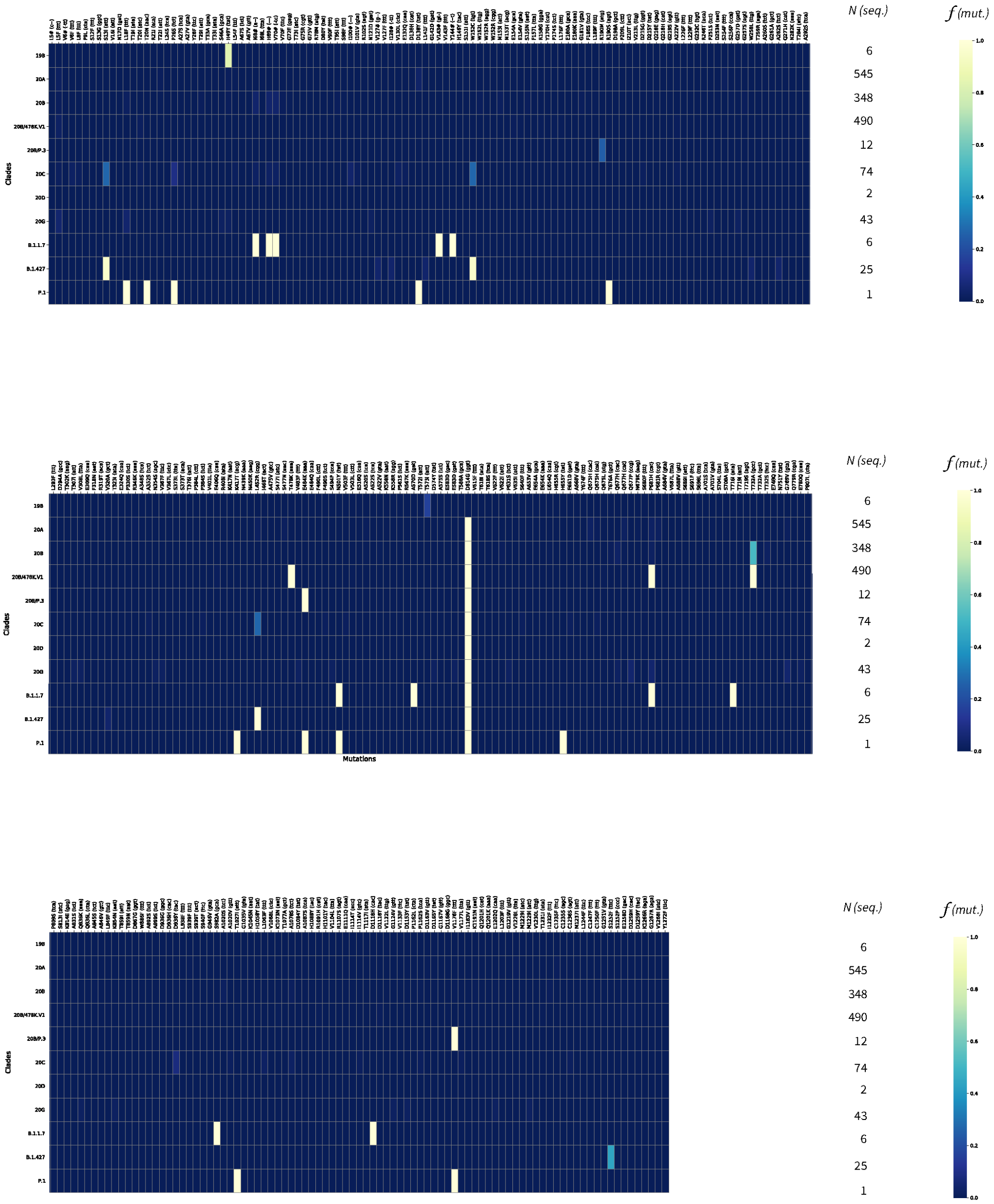


**Figure S5. Heatmap of relative frequencies (*f*) of mutations in Mexico (pre-vaccination stage).** The 315 mutations identified across 1552 viral genomes, arranged in 11 clades (19B, 20A-D, 20B/478K.V1, 20B/P.3, 20G, B.1.1.7, B.1.427, P.1), are shown. *N* indicates the number of available genome sequences per clade. (See **Figure 2A**
